# Impact of socio-economic traditions on current tobacco and tea addictions (Siberia 17^th^ to 20^th^ century)

**DOI:** 10.1101/2021.01.13.20232454

**Authors:** Matthias Macé, Camille Richeval, Liubomira Romanova, Patrice Gérard, Sylvie Duchesne, Catherine Cannet, Irina Boyarskikh, Annie Gérault, Vincent Zvénigorosky, Darya Nikolaeva, Charles Stepanoff, Delphine Allorge, Michele Debrenne, Bertrand Ludes, Anatoly Alexeev, Jean-Michel Gaulier, Eric Crubézy

**Affiliations:** International Research Laboratory “Coevolution between Human and environment in Eastern Siberia”, 37 allées Jules Guesde, F-31000 Toulouse, France and North-Eastern Federal University, 56 Belinskogo street, RU-677000 Yakutsk, Russia; CHU Lille, Unité Fonctionnelle de Toxicologie, F-59000 Lille, France; Univ. Lille, ULR 4483 – IMPECS – IMPact de l’Environnement Chimique sur la Santé, F-59000 Lille, France; Molecular Anthropology and Image Synthesis Laboratory, Université Toulouse III, 37 allées Jules Guesde, F-Toulouse 31000, France; Institute of Foreign Languages and Regional Studies, North-Eastern Federal University, 56 Belinskogo street, RU-677000 Yakutsk, Russia; Institute of Forensic Medicine, 11 rue Humann, F-67085 Strasbourg, France; Central Siberian Botanical Garden SO RAS, RU-630090 Novosibirsk, Russia; BABEL, CNRS FRE 2029, Université Paris Descartes, 12 rue de l’Ecole de Médecine, F-75006, Paris, France; École pratique des Hautes Études, Université Paris Sciences et Lettres, Les Patios Saint-Jacques 4-14 rue Ferrus, F-75014, Paris, France; Department of Romano-Germanic Philology, Institute of Humanities Novosibirsk State University, RU-630090 Novosibirk, Russia; Institute for Humanities Research and Indigenous Studies of the North (IHRISN), Petrovskogo St. 1, RU-677007 Yakutsk, Russia

## Abstract

**Objective:** To investigate how tobacco and tea spread among virgin populations and how the first addictions have subsequently influenced the behavior of present-day populations.

**Design:** Retrospective observational study using data from frozen burials and levels of theobromine, theophylline, caffeine, nicotine, and cotinine measured in hair samples from frozen bodies of autochthonous people. Confrontation of the results with new ethnobotanical, historical and cultural data from the past and with present day epidemiological data from the same region.

**Setting:** Eastern Siberia (Yakutia) from the contact with Europeans (17th century) to the assimilation of people into Russian society (19^th^ century).

**Participants:** 47 frozen bodies of autochthonous people from eastern Siberia and a review of present-day populations from Yakutia

**Intervention:** Levels of theobromine, theophylline, caffeine, nicotine, and cotinine were measured in hair samples. Along with the collection of cultural data associated with the bodies, potential comorbidities were investigated.

**Main outcome measure:** We combined LC-HRMS and LC-MS/MS tools for toxicological investigations in hair and we assessed the association between xenobiotic concentrations and geography using several permutation-based methods to infer the economic circuits of tobacco and tea. Comparison of the results obtained with ethno-botanical analyses allowed to identify the products from which the metabolites were derived.

**Results:** Hair levels of theobromine, theophylline and caffeine vary with the type of beverage consumed: green, black or local herbal teas. At the beginning of our study period, a few heavy consumers of tobacco were found among light or passive consumers. Tobacco-related co-morbidities began to be recorded one century after contact with Europeans. Heavy tea users were only found from the 19^th^ century and the heaviest users of the two substances date from this century. After the first contact, teas were widely consumed as beverages and medicines but also for shamanic reasons. Economic factors, fashion and social and family contacts seem to have played a decisive role in tobacco consumption very early on.

**Conclusion:** Epidemiological characteristics of present-day Yakutia suggest that the high prevalence of smokers and tea consumers, the prevalence of female smokers and tobacco use in the north, find their origins in the diffusion phenomena of the 18th and 19th century. Behavioral evolution governed the process of substance integration and was determinant for the continuity of use of these substances over a long period of time.

## Introduction

Understanding the historical foundations of contemporary addictions is challenging because historical data do not always reflect biological data and the latter are scarce. We had the opportunity to excavate more than 150 frozen graves in East Siberia (Yakutia) dating from the 17th to the 19th century. Some revealed smoking material another the most northerly English teapot ever found in an archaeological excavation (figure 1). We were able to find metabolites of tea and tobacco in the hair of 47 subjects. These analyzes, confronted with historical, economic and ethnological studies, reflect the way in which green tea, and later black tea and tobacco spread in Siberia and how these substances competed with native herbal teas. Passive smoking in young people and comorbidities can be described. Certain dissemination processes among these indigenous populations, either through the elites and heavy consumers or through certain distribution channels, are the same as those which today favor addictions. More curiously still, certain specificities which developed with the first consumers of the 18th century are still part of the epidemiological characteristics of modern Yakutia. This study supports public health policies based on intervening on economic circuits or modes to curb certain addictions, as it shows us that the origin of these addictions is sometimes older and more complex than one might suppose.

**Figure 1.**
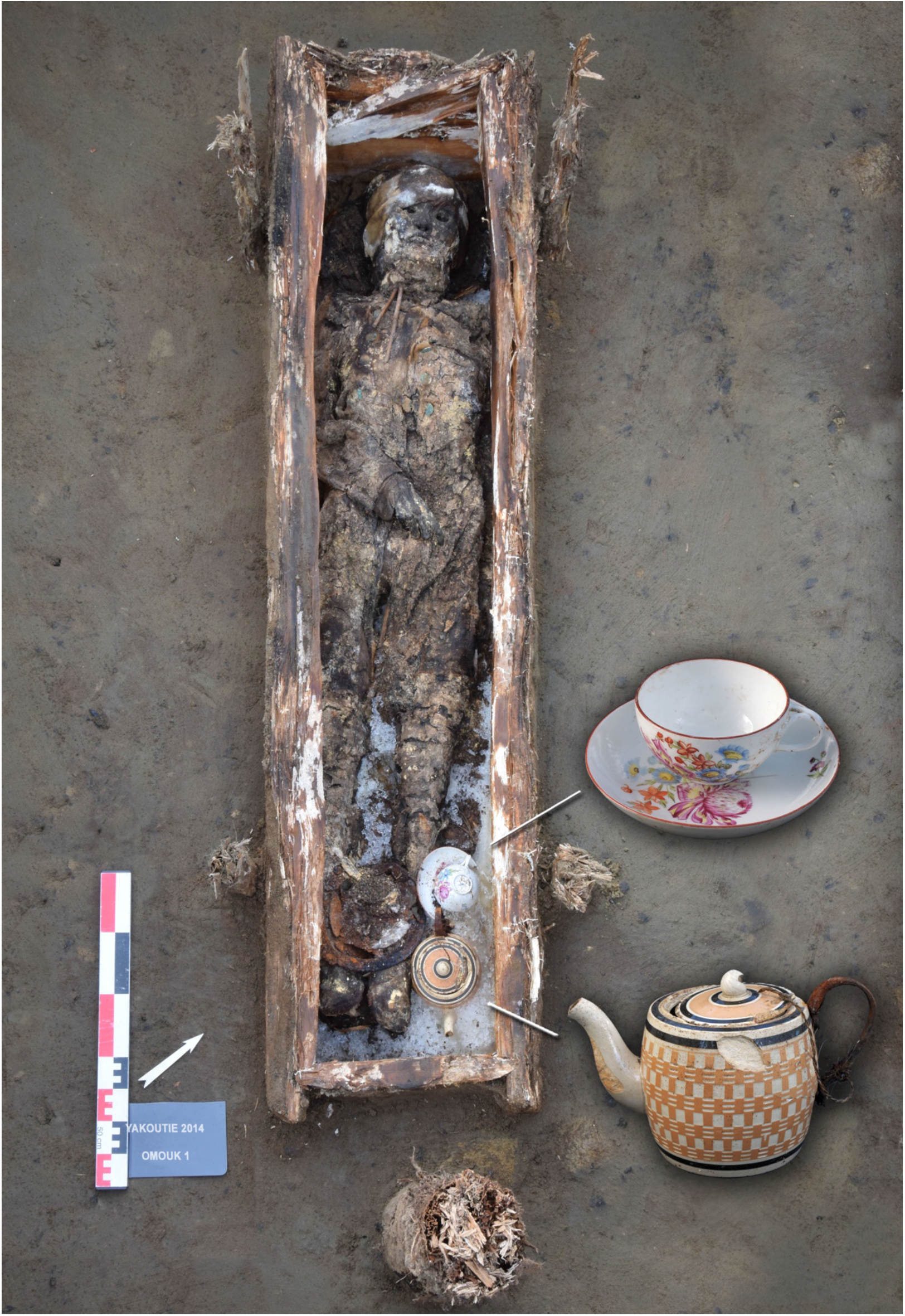
Omouk 1 grave with artefacts: teapot and cup, and under the legs and feet the smoking kit (iron lighter, fire bag, pipe). This grave (around 1820) was found on the Indigirka River, Arctic Circle. The cup originated from the Russian factory A. Popo known for its solid porcelain. The teapot is of English origin. The woman’s hair had high levels of nicotine, cotinine, and the highest concentration of caffeine, theobromine and theophylline.

## Material and Methods

More than 150 bodies found in frozen tombs four geographical regions (31,000 km^2^) of the Sakha Republic (Yakutia) were studied. They were classified into five chronological periods: before 1700, 1700-1750, 1750-1800, 1800-1850 and after 1850 AD. The first contacts of the autochthonous population with the Russians date back to 1620 AD but there was almost no trace of these contacts in material culture before 1689/1700AD. Assimilation in the Russian Orthodox culture was fully accomplished after 1800 AD.

We categorized and analyzed the characteristics of these ancient subjects and studied their familial relationships, social status and diet (SD1). Among these, 47 had well-preserved hair without contact with sediment. Hair samples were collected and stored during the autopsy of the subjects following procedures ruling out the possibility of outside contamination. They were decontaminated in the laboratory. We analyzed proximal hair segments (2 to 3 cm length) close to the skull corresponding to the last few weeks of life. Levels for the main compounds related to tea and tobacco consumption were analyzed in hair sampled from 47 frozen bodies, dated from the early contact with Europeans (1620 AD) to the assimilation of native people into Russian society (end of the 19^th^ century). In one case, remains of tea leaves were found in a teapot associated with a woman and analyzed. We were able to look for traces of pulmonary anthracosis in 14 subjects and pulmonary histological investigations for potential co- morbidities were also carried out. In 7 out of these 14 cases we were able to analyze both hair and lungs, and for one woman a breast concomitantly, to establish whether the breastfeeding woman had high nicotine use (SD2).

Previous studies investigated nicotine and caffeine in hair[1,2] and used LC-HRMS and LC-MS/MS tools for toxicological investigations in hair[3,4]. We combined these two techniques according to previously published methods[5,6] for screening drugs and toxics compounds, psychoactive drugs including natural alkaloids, and tea methylxanthines (caffeine, theobromine, theophylline) and tobacco alkaloids (nicotine) and metabolite (cotinine) (SD3). Another large toxicological screening for psychoactive drugs complied with international recommendations[7,8].

### Statistical analyses

Principal Component Analysis was performed over the levels of the five combined analytes and outliers defined qualitatively (SD4). The number of tea metabolites detected in hair varied from zero to three (theophylline, theobromine, caffeine) depending on the individual. Multivariate logistic regressions were used to assess a putative shift in the use of tobacco and tea over time (before and after 1800 AD, see SD4 for details), adjusting for gender, age, social status and presence of pipe. Spearman’s correlation coefficient between nicotine and cotinine concentrations was computed within each age class (0-15, 15-30, 30-50 & over 50 years old). Co-correlation between age classes was assessed between the age class of the children (0-15 y/o) and each of the other age classes, including Zou’s confidence intervals[9]. Computations were performed using the *Cocor* R package[10].

We assessed the association between xenobiotic concentrations and geography using several permutation-based methods (SD4). We then assessed the correlation (Spearman’s rank test) between analyte concentrations and distances from the main trading posts and/or fairs in the east through which tea and/or tobacco was likely to transit as well as the known entry areas through which it was likely to arrive (SD9). Strength of association was tested using a permutation over individual concentrations. Given the significance of the association between the levels of cotinine and/or nicotine with the pipes (SD5), we looked for the same associations, on the totality of the pipes (for a majority of subjects the hair was not preserved), before and after 1800 AD. The PCA first axis values computed as described above were interpolated over a rhomboid area comprised between the four geographical regions using spline interpolation and triangulation based on linear interpolation as implemented in the *akima* R package[11]. Maps were produced using the *marmap* R package and the NOAA bathymetrics and global relief data (resolution of 5 minutes) and then drawn following an orthographic projection for 100° longitude, 60° latitude and 0 m elevation.

## Results

Hair analyses showed a LOD of 0.01 ng/mg and LLOQ of 0.02 ng/mg for nicotine and cotinine; a LOD of 0.05 ng/mg and LLOQ of 0.01 ng/mg for caffeine, theobromine and theophylline (**Erreur ! Source du renvoi introuvable**.**Erreur ! Source du renvoi introuvable**.). It is of note that no other natural alkaloid compound or related metabolite (e.g. morphine, codeine, atropine, scopolamine, …) was detected in the 47 analyzed hair samples (SD6). The last dichloromethane bath used for the hair decontamination step tested negative for all compounds and for all samples.

Theobromine (n=31) was more often detected (including in one 6 to 9-month-old child) than caffeine (n=29), which was itself more often detected than theophylline (n=19). After 1800 AD, at least one substance was detected in all subjects, and the three substances were detected in 8/15 subjects (Table 2). The subject presenting the highest concentration was a woman buried with a teapot (SD7 and SD11). Before 1800 AD, no substance was detected in 8 hair samples and the three substances were detected in only 6/32 samples. The most frequent case (9/32) was that of subjects with hair samples containing both theobromine and caffeine, essentially women (8 females out of 9 individuals: p=0.0182) and the only male with the two substances dated from the period prior to 1700 AD (SD8). We found a sub-significant association between the status of shaman and theobromine concentrations (SD7).

**Table 1:**
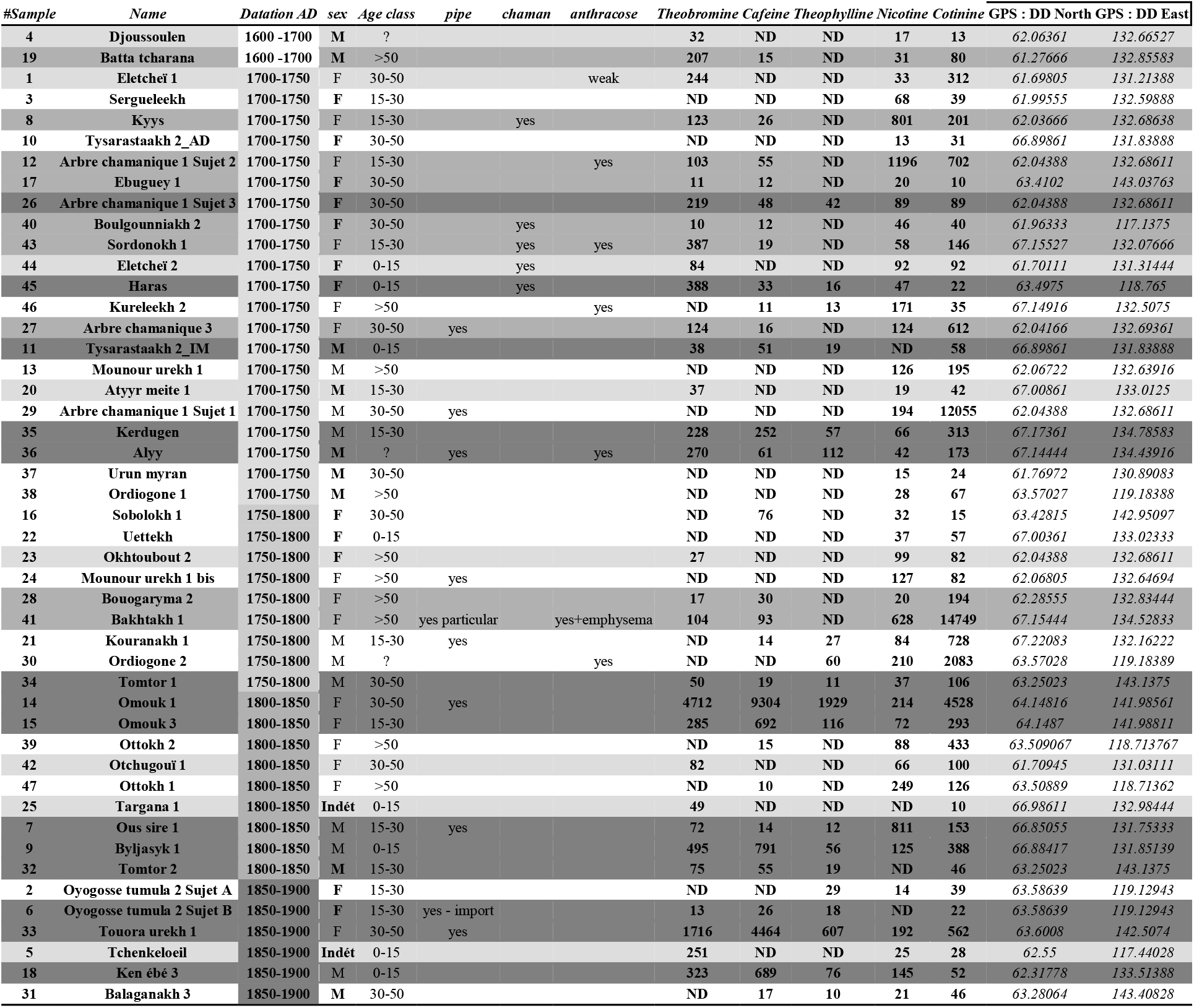
Hair levels of xenobiotics (pg/mg) by individual alongside the available datations, age class and archæological data. LOD: limit of detection, LLOQ: lower limit of quantification. The fit of the 1/x weighted quadratic regression (analyte-to-IS peak area ratio versus theoretical concentration) was verified for all the compounds. The coefficients of determination were superior to 0.99 for each calibration curve (from LLOQ to 20 ng/mg). Precision and accuracy were acceptable for all substances, meeting the criteria set: lower than 25% at LOQ and lower than 20% at other levels for both inter- and intra-day assays. The samples are listed in chronological order, from 1600 AD to 1900 AD. We note the increase of subjects presenting the three methylxanthines (caffeine, theobromine, theophylline) after 1800 AD (generalization of the black tea, dark grey), the presence of subjects with two compounds (theobromine, caffeine) between 1700 and 1800 AD (emergence of the green tea, medium grey) and the presence from 1600 AD of only theobromine (local tea: Ivan tea, light grey).

**Table 2.**
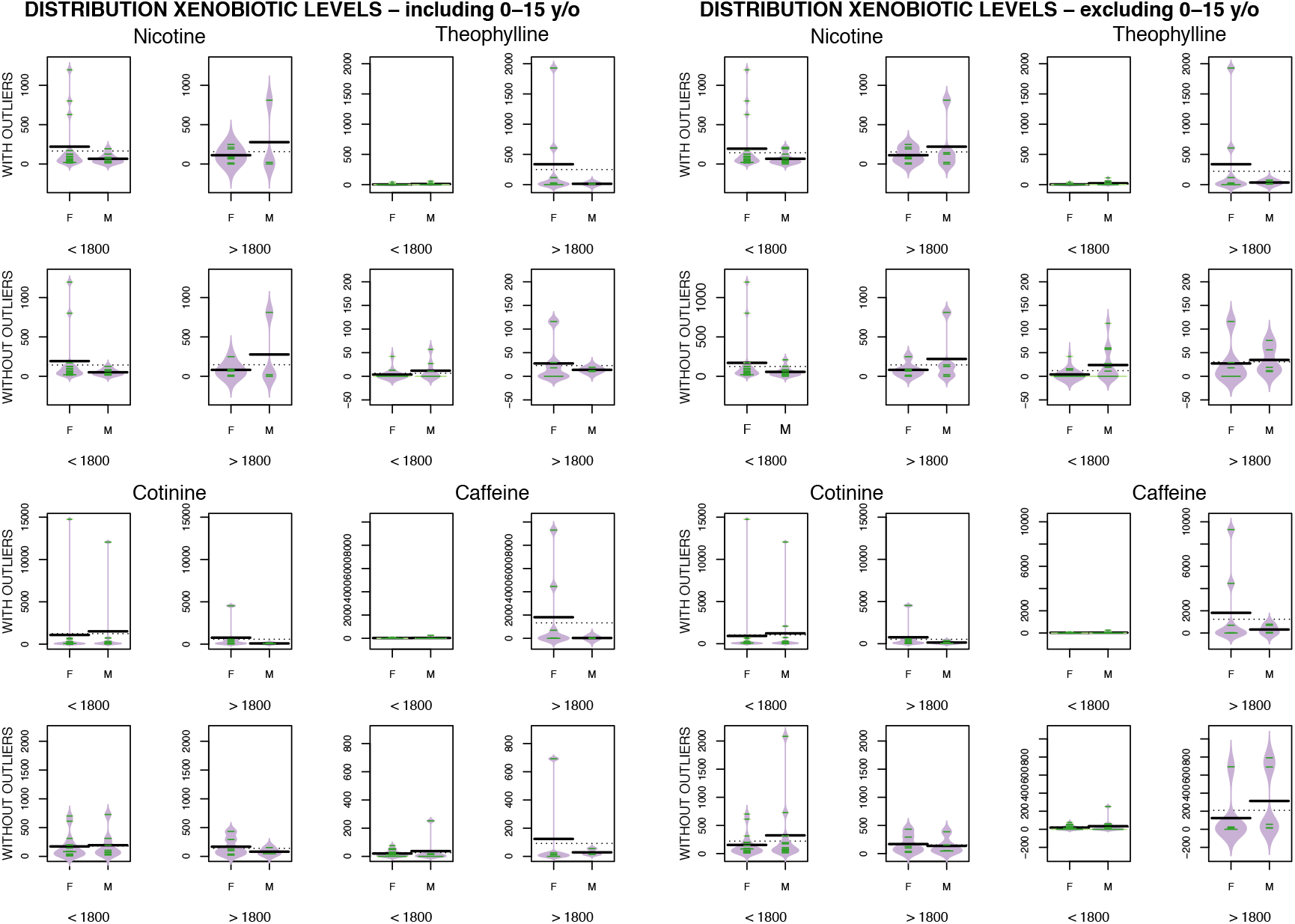
Distribution of xenobiotic hair levels over time and gender. Above: nicotine vs theophylline. Below: cotinine vs caffeine

Cotinine (n=47, 100%) was more often detected than nicotine (n=43, 91.4%). The two subjects with the lowest hair cotinine concentration were a child aged 6 to 12 months and a child aged 6 to 9 months. Cotinine was sometimes found in some adults especially those dated before 1700 AD.

The ten subjects buried with a pipe had traces and/or significant hair concentrations of cotinine and/or nicotine, one had no trace of nicotine and traces of cotinine. For graves with pipes, before 1800 AD we found a sub-significant association between the presence of pipe and the distance to Yakutsk or the distance to the south-west of the studied area (SD9). The nicotine/cotinine correlation increased with age and the correlation was significantly different between the young (0-15 years old) and the oldest quartile (over 50 years old) - SD7 -. An older woman from the end of the 18^th^ century was the only subject with a level of cotinine associated with the highest level of nicotine; anthracosis pigments found in her lungs can indicate death by -SD2-.

Eighty-two percent (39/47) of the subjects had traces of tea and tobacco in hair samples. The highest users of the two substances were from the 19^th^ century, and heavy tea users were only found in the 19^th^ century. Moderate tea users in the 19^th^ century were also tobacco users. There were families of heavy smokers and non-smokers, and in one family of heavy smokers one of the women was breastfeeding at the time of her death. In the case of caffeine and cotinine, we were able to show a negative correlation between the concentration of these substances and the distance from sampling location to the three main trading posts and/or fairs in the east through which the tea was likely to have transited (SD9). The highest correlation was with the northeast, which represents the point of entry of the trade route by which tea was brought at this period from the Pacific harbor of Okhotsk into Yakutia (p-value = 0.0112). The robustness of this association was challenged by permuting 10% of the metabolic concentration values among individuals (10,000 replicates). The association was confirmed with probability of observing r_s_ above 0.5 above 0.9 for Zachiversk harbor. This is consistent with a NE-SW gradient in caffeine concentration after 1800 AD (Figure 2).

**Figure 2.**
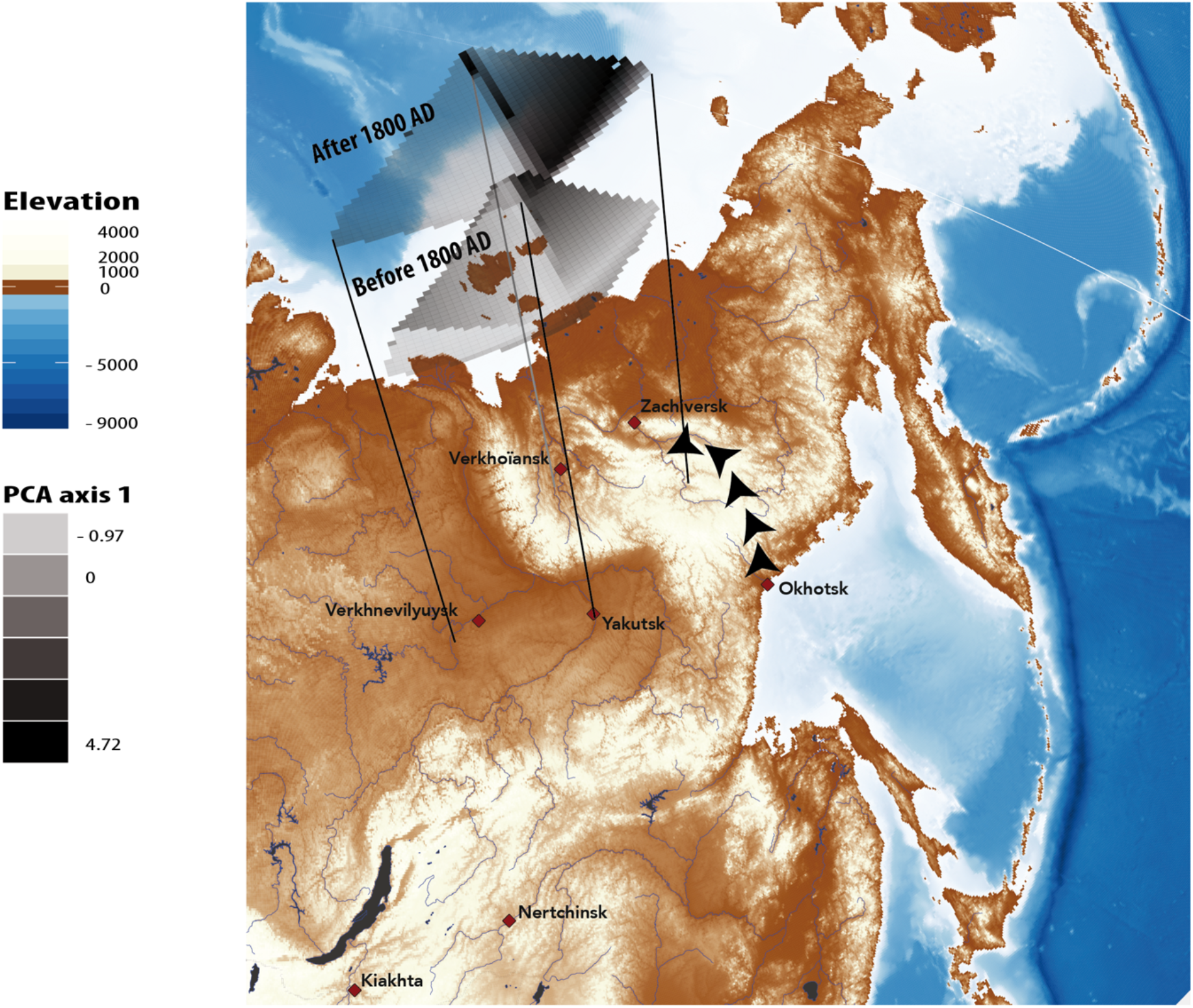
Map of Eastern Siberia displaying geographical interpolation of tea use overlaid with geographical data of the studied area. Tea use was estimated as the first axis of the Principal Component Analysis over the three studied methylxanthines (Caffeine, Theobromine, Theophylline). Eigenvalues - axis 1: 2.19; axis 2: 0.51; axis 3: 0.28. Before 1800 AD, no geographical pattern was found as only isolated individuals showed significant levels of methyxanthines associated with tea. After 1800 AD, a significant negative correlation was observed between caffeine concentrations and the settlements situated northeast (above 80% significant correlations observed after 10.000 permutations tests, see SD for details) on the path of the importations from Okhotsk harbor.

## Discussion

The use of tobacco products is the deadliest addiction worldwide and a positive association between smoking and caffeine intake is found in some populations, although the preference for the type of caffeinated drink – particularly tea versus coffee – varies greatly among countries[12]. Despite prevention campaign efforts to reduce the public health burden of tobacco, prevention has not benefitted all strata of various societies[13], because addictions involve the interplay of pharmacology, genetics, and social factors together with environmental factors, including fashion and identity[14]. Although multilevel transdisciplinary models are needed to comprehensively understand sources of tobacco-related health disparities (TRHD), the incorporation of historical context into TRHD research remains infrequent. Yakutia (north eastern Siberia) offers a unique opportunity for such research. Our study is the first to analyze such a large number of hair samples collected in the minutes or hours following the excavation of frozen bodies. Considering that the growth rate of human scalp hair ranges from 0.7 to 1.4 cm/month with a mean of about 1 cm/month[15] we analyzed samples of hair corresponding to approximately two to three months’ growth before the subject’s death, much less for the children for whom the hair was not very long. Xenobiotics are well preserved in hair[16], we were able to determine strong associations between hair concentrations of some substances and cultural factors even if exposure to second-hand smoke during the lifetime of the patients cannot be excluded[17] (SD10). The Yakuts buried their dead with items representative of their lives and/or occupations. Some strong empirical associations were found: the teapot was associated with the highest tea consumer, the heaviest smoker presented pulmonary emphysema, and the presence of a pipe was associated with smoking in 9 out of 10 cases. Cotinine/nicotine correlation increased with age, probably reflecting passive smoking in young people.

Tea contains some of the 3-methylxanthines (caffeine, theophylline and theobromine), but its chemical composition is significantly affected by tea processing, leaf maturation, botanical varieties, geographical origin, agricultural practice[18] and the way to prepare it[19]. The influence of physiological variations from one subject to another on the incorporation of metabolites in hair is, to our knowledge, unknown. However, while all three 3-methylxanthines are present in most black teas, theophylline is often absent in green teas[20] and in some black teas[21]. In our sample, women seem to have consumed green tea from the end of the 17th century. Black tea consumption, which appeared punctually before 1750 AD, became widespread in the 19th century, consistent with historical data[22]. Ivan tea, a fermented herbal tea from *Chamerion latifolium* and *Chamerion angustifolium* which was drunk at that time, contains only theobromine in quite large quantities[23]. This native herbal tea was consumed by five subjects of various ages from the 17th and 18th centuries; in the 19th century it was only detected in three subjects, two of whom were less than one year old, suggesting that at that time it could have been still used for medicinal purposes. It cannot be ruled out that other herbal teas may have been consumed. (SD11).

All periods combined, we found a few heavy tobacco consumers, men and women, amongst light or passive consumers, but tobacco-related co-morbidity was only recorded after 1750 AD (in a woman) at a time when a large part of the population, even older children, became addicted according to travelers’ accounts[24,25]. Heavy tea users were only detected in the 19^th^ century and the heaviest users of the two substances dated from this century. In the 19th century, while the Yakut population assimilated into the Russian orthodox way of life, several contemporary trends were detected: all subjects seemed to become, at least, light consumers of tea, and moderate tea users were also tobacco users. The average tobacco-related xenobiotic hair concentration was higher in women than in men before 1800 AD (Figure 4) and this peculiarity is still found today in the Sakha republic and the current population of the Republic currently exhibits generalized tea drinking habits and a higher smoking incidence than any other part of the Russian Federation[26,27]. As far as the further away from Verkhoyansk (in the north), the lower the nicotine concentration in hair. This probably established the current feature of the prevalence of smokers in the north[26].

The generalization of tea and tobacco is linked to economic circuits. In our study area, tobacco use spread faster than tea use, probably because tobacco is intrinsically more addictive than tea, and because of the absence of a previous tradition in the use of these substances. A variety of herbal teas -specially Ivan tea- have been used by the Yakuts for centuries. Herbal teas were widely consumed as beverages and medicines for disease prevention and treatment but also for shamanic reasons as attested by the theobromine levels in female shamans. The use of green tea by women in the 18^th^ century also support this hypothesis, and the generalization of the use of tea follows the development of tuberculosis[28] for which green tea was used as a treatment, perhaps with some results[29]. In the 19^th^ century, tea had made the transition from a beverage used in medicine and spirituality-based socialization, to being used by elites with the tea ceremony led by women of fashion and means. The ceremony dictating that tea must be brewed and drunk using specialized equipment (**Erreur ! Source du renvoi introuvable**.) soon became established as a more generalized practice. For tobacco there were no previous traditions; socio-economic and family contacts seem to have played a decisive role very early with this substance, which is more addictive than tea.

The earliest subject (#4) presented only a few traces of cotinine and nicotine in hair, perhaps reflecting passive exposure due to contact with Europeans as some of his artefacts demonstrate. Another man (#19) with slightly higher concentrations of these substances died at a time when a 1680 report from the European mercenaries underlined that they traded tobacco for furs with autochthones[30]. Less than a century after contact, proximity to the major tobacco import trading post of Kiakhta was a determining factor in consumption as were direct relationships of individuals with this spot. The development of economic circuits affected consumption, which largely flouted laws - as early as 1634 AD, smoking was prohibited in Russia on punishment of death (1634) and it remained so for autochthonous people until 1822 AD. The development of smoking was linked to the economic circuits in the 18^th^ century, while in the 19^th^ century tobacco consumption became generalized and economic circuits were detectable only for tea which was imported via the Pacific harbors. Between 1700 AD and 1800 AD two men of the elite (#29 and #30) with a Mongolian haircut and a southern fashion for their clothes were heavy consumers of tobacco. One of them was found with a beautiful mammoth ivory pipe. The manufacturing of highly attractive smoking accessories was instrumental in the dissemination of tobacco (SD 5). The custom then was to pass the pipe from one person to the other to smoke thus encouraging family and matrilineal transmission. These men probably travelled to a trading post on the Mongolian border in contact with China where tobacco, tea and furs were traded. Two other contemporary heavy smokers, females in this case (#12 and #26), were related to one of them and one was breastfeeding when she died. The mother of #29 and #30, who was not supposed to be living under the same roof as them when she died, was not a heavy tobacco user and drank a different tea than her daughter. Similar family habits were also observed in the 19^th^ century.

## Conclusions

After the first contact, teas were widely consumed as beverages and medicines but also for shamanic reasons; for tobacco, socio-economic factors, fashion and family contacts seem to have played a decisive role very early on. It seems therefore that behavioral evolution governed the process of integration of tobacco and tea into the Yakut culture, and in so doing, led to the continuity of use of these substances over a long period of time. Some epidemiological characteristics of present-day Yakutia find their origins in the diffusion phenomena of the 18th and 19th century. Analyzing the respective contributions of social and economic processes in the use of these substances opens avenues of investigation for today’s public health.

## Supporting information

https://we.tl/t-xk1bnImUfr

## Data Availability

On request all the data provided in this article are available from the corresponding authors.

## Compliance with Ethical Standards

### Funding

This work was supported by the program of the France-Russia Associated International Laboratory (LIA COSIE number 1029), associating the North-Eastern Federal University (Yakutsk, Sakha Republic), the State Medical University of Krasnoyarsk, the Russian Foundation for Fundamental Research (Moscow, Russia), the University of Paul Sabatier Toulouse III, the University of Strasbourg I (France) and the National Centre for Scientific Research (Paris, France). Funding for excavations was provided by the French Polar Institute Paul Emile Victor, the French Archaeological Mission in Oriental Siberia (Ministry of Foreign and European Affairs, France), the North-Eastern Federal University (Yakutsk, Sakha Republic).

### Ethical approval

this article does not contain any studies with human participants or animals that were performed by any of the authors.

### Informed consent

not applicable *[Kaufmann IM, Rühli FJ. Without ’informed consent’? Ethics and ancient mummy research. J Med Ethics. 2010;36(10):608-13. doi: 10.1136/jme.2010.036608]*

