## Supplementary material for "Impact of socio-economic traditions on current tobacco and tea addictions (Siberia 17^th^ to 20^th^ century)": https://we.tl/t-xk1bnImUfr

Matthias Macé, MVDr, PhD,
Camille Richeval, PhD,
Liubomira Romanova, M.Sc.,
Patrice Gérard, M.Sc.,
Sylvie Duchesne, M.Sc.,
Catherine Cannet,
Irina Boyarskikh, PhD,
Annie Gérault, MD,
Vincent Zvénigorosky, PhD,
Darya Nikolaeva,
Charles Stepanoff, PhD,
Delphine Allorge, Pharm D, PhD,
Michele Debrenne, PhD,
Bertrand Ludes, MD, PhD,
Anatoly Alexeev, PhD,
Jean-Michel Gaulier, PharmD, PhD,
Eric Crubézy, MD, PhD,, +33 6 11 60 67 89

### Supplementary data

#### Tables

#### Figures

#### SD1: Study design and data collection

During the past 18 years, we have surveyed 31,000 $km^2$ of particularly suitable landscape in the Sakha Republic (Yakutia) and completed the archaeological study of more than 150 bodies (some still frozen) dating from the 15th to the 19th century. For many subjects we also have indications of social status including artefacts found in tombs[1].

The samples were collected and stored by forensic scientists and professional paleopathologists during the autopsy of the subjects, within minutes or hours of the opening of the tomb, avoiding the possibility of outside contamination. Out of the 47 subjects, 10 subjects were found with a pipe, and a teapot and a teacup were associated with one early 19th century subject. Remains of brewed tea were found in the latter. In order to interpret the results, we conducted an ethno-pharmacological and historical study to determine what substances may have been consumed by the Yakuts at different periods.

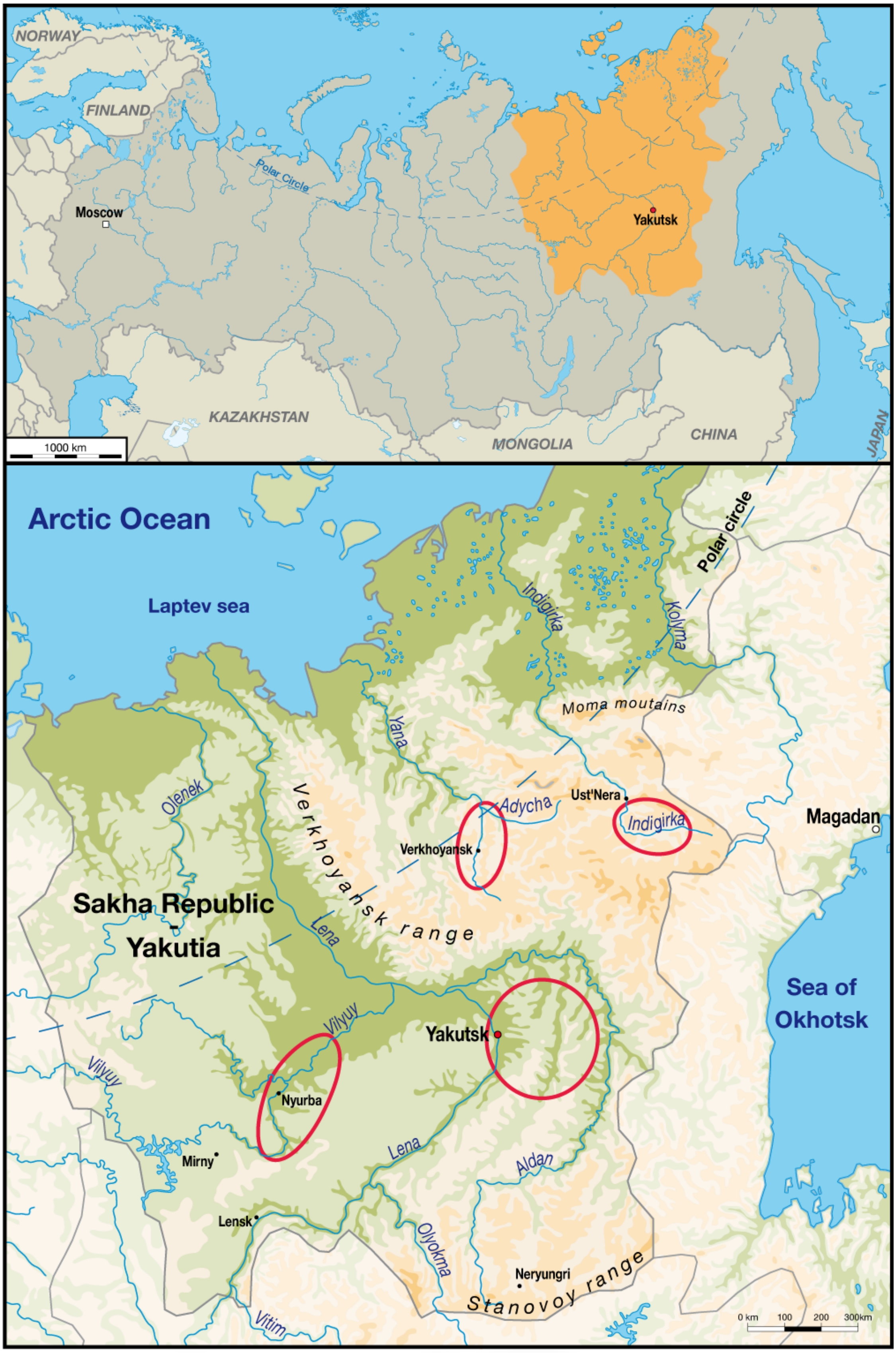

*Figure S 1. Map of Yakutia showing the location of excavation sites related to regions originally inhabited by Yakut people.*

#### SD2: Lungs and Left breast sample preparation and analysis

##### Lungs and Left breast sample preparation

From our sample of 150 bodies we were able to find traces of pulmonary anthracosis in 14 subjects. In 7 out of these 14 cases we were able to study hair and lungs concomitantly to establish whether high nicotine use was associated with anthracosis. In one case, the left breast of a woman with hair (shaman tree, number 2) was well preserved and presented high levels of cotinine and nicotine. One of the children -deceased between one and three years-, buried in the same grave as her, was her son[2], so we check whether she was breastfeeding at the time of death.

***Lungs*** were rehydrated in Ruffer I solution for 3 hours and then immersed in 10 % neutral buffered formalin for 7 days. After fixation, tissues were dehydrated through increasing grades of ethylic alcohol, cleared in xylene and embedded in paraffin wax. Tissue sections of 5 µm thickness were stained with hematoxylin and eosin (H&E) to assess the general morphology. For one woman, associated with her young son (whose hair wasn't preserved) in a mass grave we could study hair and left breast to determine whether doses of nicotine were associated with breastfeeding.

***Left breast :*** Tissues were naturally mummified by freezing and remained in this state until they were removed from the funeral chamber and autopsied. Upon reception in the laboratory tissues were rehydrated, given their very dry aspect, before the histological processing. Tissues were rehydrated in Ruffer I solution[3]. Rehydration was completed after 3 h.

Once rehydrated, tissues were fixed in 10% buffered formalin for 4 days. One third of the tissues was deep frozen and the remaining pieces dehydrated through increasing grades of ethylic alcohol, cleared in xylene and embedded in paraffin wax.

Serial sections of 4 µm in thickness were cut and stained with: (i) hematoxylin and eosin (H&E) to assess the general morphology on paraffin sections, (ii) Giemsa stain for the demonstration of parasites on paraffin sections, (iii) Sudan III and IV for the demonstration of lipids on frozen sections.

##### Lungs analysis: Anthracosis and emphysema

Eight lungs out of 14 presented traces of anthracosis (see Figure S 2). There is no relationship with sex (five women, three men), nor with age (15-year old subjects presented traces, some over 60 didn't). Two subjects with very low levels of nicotine didn't show signs of anthracosis, while in two other subjects, nicotine levels and presence/absence of anthracosis were in opposite directions. Bakhtahh 1, an older woman who was found with a unique model of pipe, was the only case with level of cotinine associated with the highest level of nicotine; she died with anthracosis pigments associated with emphysema.

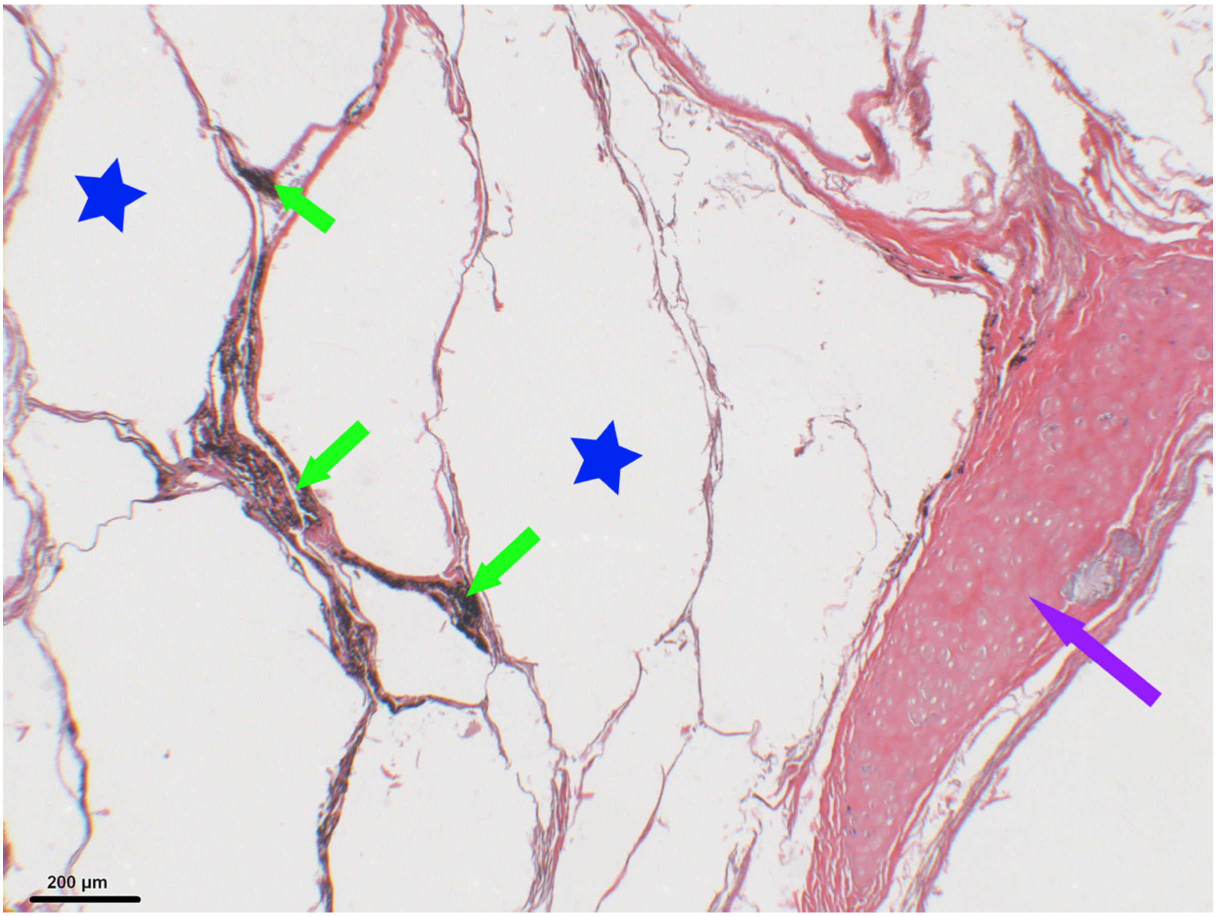

*Figure S 2. Histological slice of Baktakh_1 right lung (old woman). Anthracosis is shown by green arrows, associated emphysema by blue stars et bronchus hyaline cartilage by the purple arrow.*

##### Histology of the left breast

At histology, the lobular aspect was clearly identifiable and the lobules were separated by connective fibers (Figure S 3). Numerous secretory acini were found within a fine collagenous stroma (Figure S 4). The positive Sudan III and IV stain, confirmed the presence of lipids within the acini (Figure S 5, Figure S 6).

The abundant alveolar aspect of the gland and the scarcity of the collagen fibers are suggestive of a lactating or a pregnant breast.

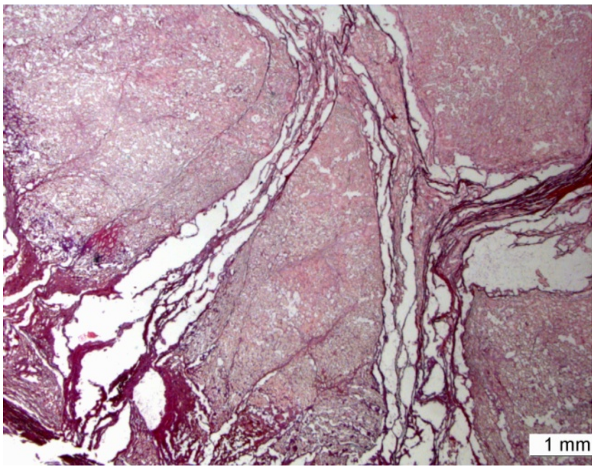

Figure S 3. Lobular architecture and connective fibers.

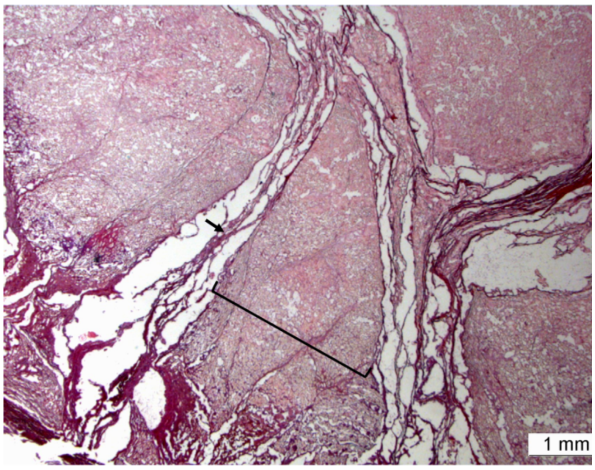

Figure S 4. Lobular architecture (bracket) and connective fibers (arrow).

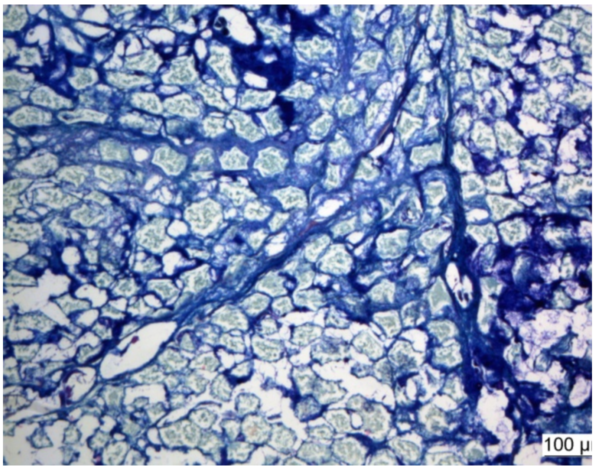

Figure S 5. Left breast (Giemsa). Numerous secretory acini within fine collagenous stroma.

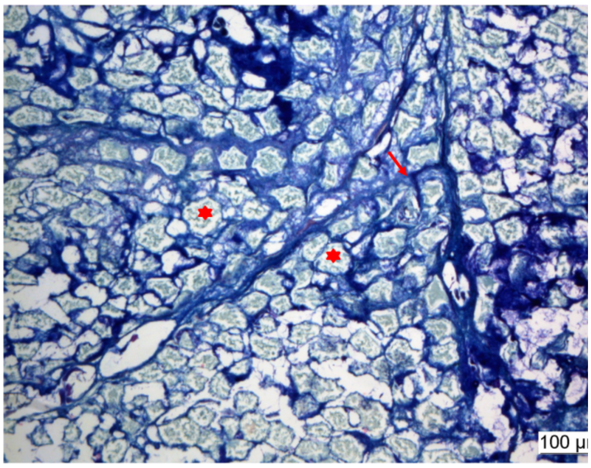

Figure S 6. Left breast (Giemsa). Numerous secretory acini (star) within fine collagenous stroma (arrow).

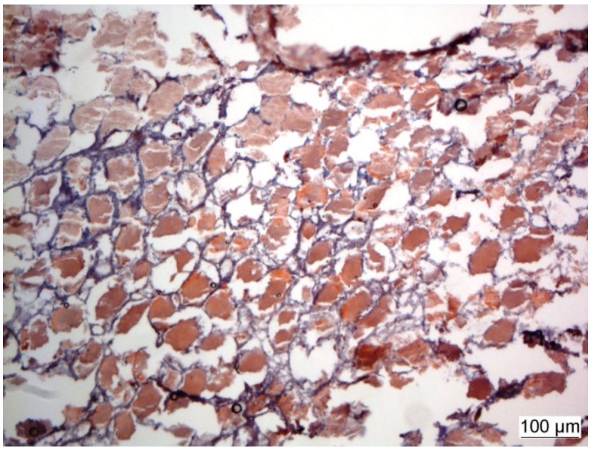

*Figure S 7. Left breast (Sudan III and IV). Lipid contents within acini.*

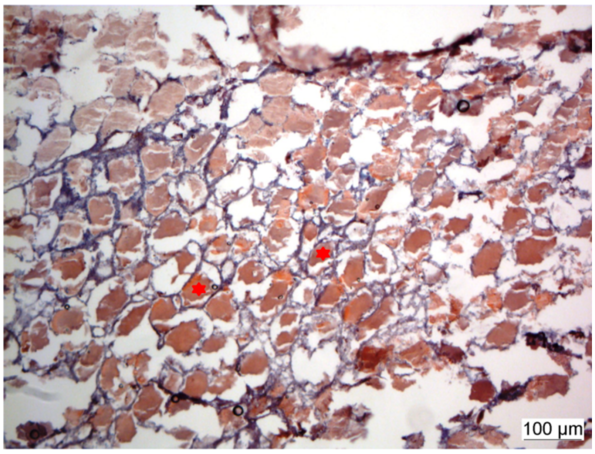

*Figure S 8. Left breast (Sudan III and IV). Lipid contents within acini (star).*

#### SD3 Methods for the detection of tea methylxanthines (caffeine, theobromine, theophylline) and tobacco alkaloids (nicotine) and metabolite (cotinine), and drugs and toxics compounds.

##### Hair samples preparation and analysis

Hair samples (n=47) were photographed and decontaminated in 2 x 2 min baths in water, followed by 2 x 1 min baths in dichloromethane. After drying, proximal hair segments (2 to 3 cm length) were cut into small pieces (<1 mm length). For extraction, 20 mg of hair samples were incubated in methanol and the solution obtained after centrifugation was used for analysis. Previous studies on nicotine and caffeine analyses in hair[4–6] and the use of LC-HRMS and LC-MS/MS tools for toxicological investigations in hair [7,8] supported our decision to use these two analytical methods for the detection of tobacco and tea products in our ancient population sample. We combined these two techniques according to previously published methods[9–11] for screening drugs and toxics compounds, psychoactive drugs including natural alkaloids, and tea methylxanthines (caffeine, theobromine, theophylline) and tobacco alkaloids (nicotine) and metabolite (cotinine).

1 - A large screening for drugs and toxics compounds was performed in hair samples using LC-HRMS. We used a Waters ultra-performance liquid chromatography system consisting of two binary solvent manager LC pumps, a sample manager autosampler and an Acquity column manager oven. Mass spectrometry data were acquired using a XEVO G2-XS QTOF instrument (Waters, Manchester, UK) controlled with MassLynx 4.1 software. After addition of 50 µL of internal standards (methyl-clonazepam and β-OH-ethyltheophyllin) to 100 µL of the methanolic hair extract, the obtained mixture was subsequently evaporated to dryness at + 30 °C under a gentle stream of nitrogen. The dry residue was reconstituted using 50 µL of a mixture of ammonium formate buffer 5mM, pH 3/acetonitrile in 1 % formic acid (87/13 ; v/v): 10 µL were injected in the chromatographic system. Chromatographic separation was performed using an ACQUITY HSS C18 column (150 x 2.1 mm, 1.8 µm, Waters) in an oven temperature of 50 °C, and mobile phases including ammonium formate buffer 5mM at pH 3 and acetonitrile in 0.1 % formic acid. For detection, mass spectrometric conditions were as follows: positive electrospray ionization interface (ESI+), ion spray voltage set at 20 V, source temperature set at 140 °C and desolvation temperature at 500 °C with a desolvation gas flow rate of 900 L/h, nitrogen as desolvation gas and argon as collision gas. The data were processed using ChromaLynx, TargetLynx, MassFragment and MetaboLynx associated software (Waters) using a homemade database of more than 1,400 substances.

2 - Using LC-MS/MS (XEVO TQ-S system, Waters, Manchester, UK), another screening for psychoactive drugs including natural alkaloid compounds and related metabolites was performed in hair samples. After addition of 50 µL of internal standards (methyl-clonazepam and β-OH-ethyltheophyllin) to 50 µL of the methanolic hair extract, the obtained mixture was subsequently evaporated to dryness at + 30 °C under a gentle stream of nitrogen. The dry residue was reconstituted using 100 µL of ammonium formate buffer 5mM at pH 3: 10 µL were injected for separation using an Acquity UPLC HSS C18 column (150 x 2.1 mm, 1.8 µm, Waters) and a gradient including ammonium formate buffer 5mM at pH 3 and acetonitrile in 0.1 % formic acid. Xevo TQ-S tandem mass spectrometer was used for detection after positive electrospray ionization mode in the MRM mode (using two transitions for each analyte) for more than 300 compounds and metabolites.

3 - Lastly, detection in hair of caffeine, theobromine, theophylline, nicotine and cotinine was performed using the same LC-MS/MS device (XEVO TQ-S system, Waters, Manchester,

UK). Calibration curves were extemporaneously achieved in the 0.1 to 20 ng/mg range for caffeine, theobromine and theophylline, and in the 0.2 to 20 ng/mg range for nicotine and cotinine using a blank head hair sample. After addition of 50 µL of internal standards (nicotine-D4 and cotinine-D3 at 5 µg/L) to 100 µL of the methanolic hair extract, the obtained mixture was subsequently evaporated to dryness at + 30 °C under a gentle stream of nitrogen. The dry residue was reconstituted using 100 µL of ammonium formate buffer 5mM at pH 3. Chromatographic separation of this extract was performed using an Acquity UPLC HSS C18 column (150 x 2.1 mm, 1.8 µm, Waters) and a gradient of (A) ammonium formate buffer 5mM at pH 3, and (B) ACN / 0.1 % formic acid as mobile phase at a flow-rate of 0.4 mL/min during 5 min. The oven temperature was set at 50°C and the injection volume was 10 µL. The gradient elution started with 100 % of A and decreased to 50 % at 3 min, and to 5 % at 3.3 min. The washing step with 95 % of solution B was held from 3.3 min to 4 min and the initial condition was applied from 4.1 min to 5 min. Xevo TQ-S tandem mass spectrometer was used for detection after positive electrospray ionization mode in the MRM mode using the following transitions: m/z 195.2 to 138.0 (for quantitation) and m/z 195.2 to 110.0 for caffeine; m/z 181.1 to 68.9 (for quantitation) and m/z 181.1 to 95.9 for theobromine; m/z 181.1 to 124.0 (for quantitation) and m/z 181.1 to 68.9 for theophylline; m/z 163.2 to 132.0 (for quantitation) and m/z 163.2 to 117.0 for nicotine; m/z 177.1 to 79.9 (for quantitation) and m/z 177.1 to 98.0 for cotinine; m/z 167.2 to 136.0 (for quantitation) and m/z 167.2 to 134.1 for nicotine-D4; m/z 180.1 to 80.0 (for quantitation) and m/z 180.1 to 100.9 for cotinine-D3. The validation procedure applied for this analytical method complied with both the French Analytical Toxicology Society (SFTA) and international recommendations for the validation of new analytical methods[12,13] including linearity, limit of detection (LOD), lower limit of quantification (LLOQ), accuracy, precision and matrix effects. Calibration curves, estimated using 1/x weighted quadratic regression, were considered acceptable if the coefficient of determination ($r^2$) was at least 0.99. The LLOQ was the lowest concentration with the two transitions presence and an intra-assay precision CV% and a relative bias lower than 25%. Intra-day and inter-day precision were calculated by analyzing 3 concentration levels in five replicates on five different days. Relative standard deviation (RSD) and percentage deviation of the average concentration from the corresponding nominal value were used to estimate precision and accuracy, respectively. These parameters were considered acceptable when they were lower than 25 % at the LLOQ, and lower than 20% for other levels. Ion suppression phenomenon was studied following the experimental system previously proposed[14]. Briefly, a standard solution containing the compounds of interest (at 100 µg/L) was continuously and directly infused into the mass spectrometer interface. A simultaneous LC flow containing either a pure mobile phase or a blank biological extract (blank hair) was introduced through a tee. Evolution of the signal of the transitions at the retention times of the corresponding compounds of interest was studied to determine the presence and intensity of ion suppression.

##### Chemical and reagents

β-OH-ethyltheophyllin, methyl-clonazepam, 5-sulfosalicylic acid, ammonium formate and formic acid, were purchased from Sigma-Aldrich (Saint-Quentin-Fallavier, France). LC-MS grade water and acetonitrile were purchased from Biosolve (Dieuze, France), while acetonitrile and methanol HPLC grade and 30 % hydrochloric acid were purchased from VWR Prolabo (Fontenay-sous-Bois, France).

#### SD4 Statistical Analyses

Principal Component Analysis was computed over the levels of the five pooled hair analytes, and outliers defined qualitatively on the PCA projection.

##### Logistic Regression for hair concentration throughout time

Multivariate logistic-regression analysis was used to assess a putative shift in the use of tobacco and tea over time. The dependent variable was binary coded: 0 for individuals dated before 1800, 1 for individuals dated after 1800. The independent variables were the hair concentrations in the five studied xenobiotics and potential confounding factors that were adjusted for in the multivariable analyses included sex, age (0 to 14, 15 to 29, 30 to 49, > 50), social status (suspected shaman or not), presence of an associated pipe (absence, presence: simple one, special one and imported one). Computations were made on data including and excluding outliers.

##### Association between the number of metabolites and time

The number of tea metabolites detected in hair varied from 0 to three (theophylline, theobromine, caffeine) depending on the subject. We tested the variations over time (before and after 1800) for three possible combinations: theobromine alone versus all others, theobromine+caffeine versus theobromine+caffeine+theophylline, all three metabolites versus nothing.

##### Correlations Nicotine/Cotinine among age classes throughout the time range

*Correlation by age class***:** Spearman correlation coefficient was computed between nicotine and cotinine concentrations within each age class (0-15, 15-30, 30-50 & over 50 years old).

*Co-correlation between age classes:* Co-correlation between age classes was assessed between the children age class (0-15 y/o) and each other age class. The computation was performed with data including and not including the 4 outliers (#14, 29, 31, 41, Figure S 10). One-sided Fischer Z test was conducted, with the null hypothesis being that correlation is lower in the children group than in the adult group, and the alternative being that children data display a lower correlation coefficient. Zou's confidence intervals[15] was also computed. Computations were made using Cocor R package[16].

##### *Geographic variations of xenobiotic concentrations and pipes*

Association between xenobiotic concentrations and geography was assessed using several permutation-based methods. First a permutation ANOVA was performed using the four archeological fields (North, East, West and South) as groups[17]. We then assessed the correlation (Spearman's rank test) between analyte concentrations and distances from the three main trading posts and/or fairs in the east through which the produce was likely to transit. Then, we assessed the correlation with the known entry areas (North-East and East) through which the substance was likely to arrive if it had been shipped to one of the ports on the Pacific coast (Okhotsk or Ayan) or the west, shouth-east and north-west routes of arrival in Yakutia of products from the southern trading posts (on the Mongolian or Chinese border, Kiakhta or Nertchinsk) and/or from Russia (Verkhnevilyuysk). Strength of association was tested using a permutation over individual concentration.

As the associations between the levels of cotinine and/or nicotine with the pipes were statistically significant (9/10), we carried out the same methods of association on the totality of the pipes (for a majority of subjects the hair was not preserved), before and after 1800 AD. We present only the result before 1800AD (20 pipes), the only positive one.

##### Interpolation & mapping of PCA by product

Tea and tobacco use were then approximated by combining the correspondent chemical xenobiotics by PCA (methylxanthines for tea, nicotinoids for tobacco). Resulting PCA first axis values were interpolated over a rhomboid area comprised between the four studied archaeological fields using spline interpolation and triangulation based on Renkas tripack [18] for linear interpolation as implemented in the “akima” R package. The interpolated values were overlaid according to their geographic coordinates over the map as black transparency (values normalized between 0 and 0.7). Maps were produced using the “marmap” R package and the National Oceanic and Atmospheric Administration bathymetrics and global relief data (resolution of 5 minutes) and then drawn following an orthographic projection for 100° longitude, 60° latitude and 0 m elevation.

#### SD5 Types of pipes associated with frozen bodies and epidemiological and/or cultural implications.

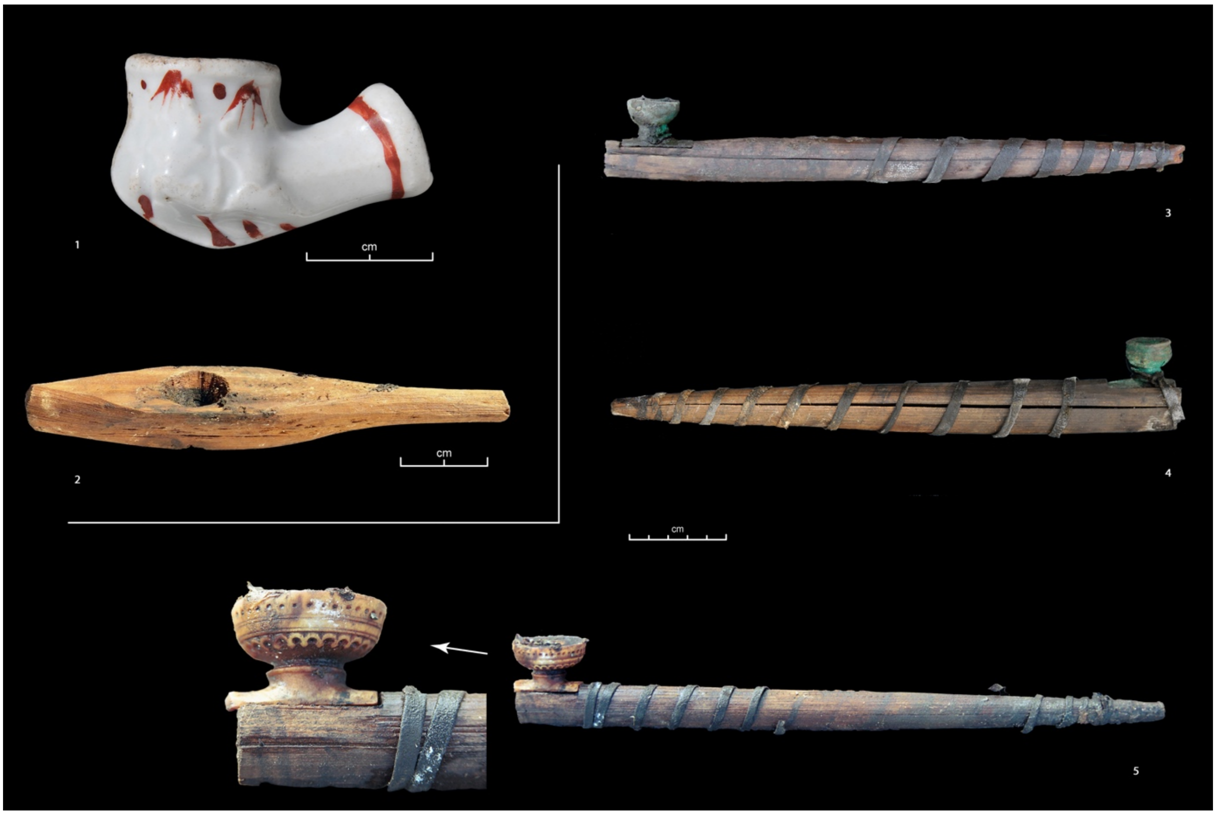

*Figure S 9. Pipes.*

(1) Pipe bowl of European origin in white porcelain - single case. Grave of a female subject aged 25 to 30 years (Oyogosse tumula 2), without any trace of cotinine or nicotine in the hair. The bowl, found without a pipe, was perhaps more an ostentatious artefact linked to the spread of tobacco fashion in the early 19th century than a useful object for smoking.

(2) Wooden pipe of a unique design that evokes a *chillum*. It was found tied to a silk scarf placed in the left hand of Bakhtakh 1 (#41 a woman from the second half of the 18th century). This woman had the highest hair levels of nicotine and cotinine in the sample and suffered from pulmonary emphysema. One instance of chillum smoking leads to maximal increase in eCO levels indicating the possibility of chillum being the most dangerous mode of smoking[19].

(3 et 4) Common pipe models. The pipe, made of wood, is kept closed by a leather lace and the bowl, made of copper alloy, is an imported good, resulting from trade and exchange with Europeans. These smoking accessories, produced in large quantities and easily accessible, increased tobacco consumption while meeting the demand. (3) Pipe found in the multiple tomb of Shamanic Tree 1 (Central Yakutia region, first half of the 18th century), slipped into the right boot of one of the two female subjects, the eldest (30 to 55 years old) and the mother of the other members buried in this tomb (her daughter, her son and two grandsons, children of her daughter). (4) Pipe found in the grave of a teenager aged between 15 and 18, at the Kuranakh site (Verkhoyansk region, second half of the 18th century), accompanied by an iron lighter.

(5) Mammoth ivory pipe, finely carved of native manufacture, found in the multiple tomb of Shamanic Tree 1 (cf. No. 3) associated with a man between 30 and 50 years old. He was a heavy smoker, belonging to the elite who was in contact with the Kiakhta trade post where the exchange of tea and tobacco took place. This type of pipe, of native manufacture and carried by the elite was an attractive smoking accessory which instrumented the dissemination of tobacco among the less privileged who bought manufactured pipe bowls (3 and 4).

#### SD 6 Caffeine, theobromine, theophylline, nicotine and cotinine hair levels

Hair analysis showed a LOD of 0.01 ng/mg and LLOQ of 0.02 ng/mg for nicotine and cotinine; a LOD of 0.05 ng/mg and LLOQ of 0.01 ng/mg for caffeine, theobromine and theophylline (Table 1 in paper). No others drugs or toxics compounds (including natural alkaloid compounds) were detected in the 47 hair samples. The second dichloromethane bath tested negative for all compounds and for all samples.

Theobromine (n=31) was more often detected (including in one child of 6 to 9-month-old) than caffeine (n=29), which was itself more often detected than theophylline (n=19). After 1800 AD, all subjects had at least one substance, and 8/15 had all three. The subject presenting the highest concentration was a woman buried with a teapot. The remains of tea leaf from the teapot tested positive for caffeine (+++), theophylline (++) and theobromine (+), whereas her hair tested positive for these three substances but with a concentration of theobromine higher than that of theophylline. Before 1800 AD, in 8 subjects no substance was detected and only 6/32 had all three; the most frequent case (9/32) was that of subjects containing both theobromine and caffeine, essentially women (8/9: p=0,0182 for comparison between sex). The only male tested positive was from the period prior to 1700 AD. There is a sub significant association between shaman status and theobromine concentration.

Cotinine (n=47, 100%) was more often detected than nicotine (n=43, 91.4%). One of the two subjects with the lowest level of cotinine was a child aged 6 to 12 months and a child aged 6 to 9 months (detection at very low levels). Cotinine was sometimes found in some adults especially those before 1700 AD.
The ten subjects buried with a pipe had traces and/or significant concentrations of cotinine and/or nicotine, one had no trace of nicotine and small traces of cotinine. Comparison of correlation (co-correlation) between age classes are given (table 3) showing an increase of the correlation with age. The difference was significant between the young (0-15 years old) and the oldest quartile (over 50 years old).

An older woman from the end of the 18$^{th}$ century was the only case with level of cotinine associated with the highest level of nicotine; she died with anthracosis pigments associated with emphysema.

Eighty-two percent (39/47) of the subjects had traces of tea and tobacco. The highest users of the two substances were from the 19$^{th}$ century, and high tea users were only found in the 19$^{th}$ century. Moderate tea users in the 19$^{th}$ century were also tobacco users.

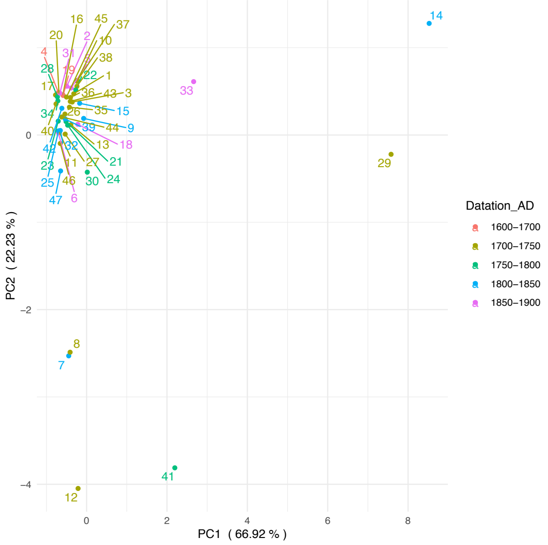

1.

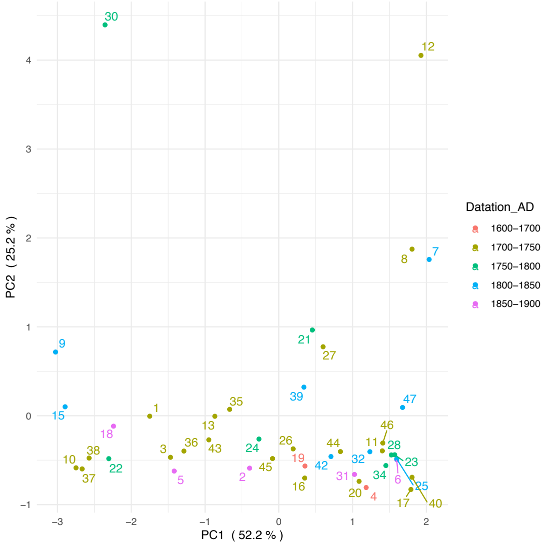

2.

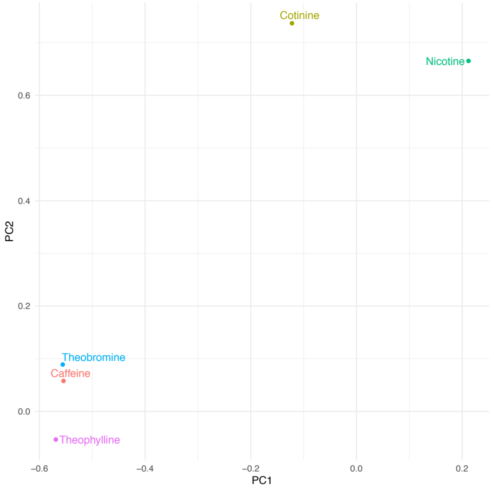

3.

*Figure S 10. Principal Component Analysis on xenobiotic concentrations.*

1. *PCA with outliers*

   *Four outliers were identified. Two (before 1800 AD) had huge hair levels of cotinine, one after 1800 AD had huge hair levels of cotinine and tea compounds, one just before 1900 AD had huge levels of tea compounds.*

2. *PCA without outliers*

3. *Contribution of each xenobiotic to PCA classification (without outliers)*

#### SD 7 Association to life history parameters

##### Samples and kinship

Hair analyses were possible in almost a third of the excavated subjects (the best preserved by the freezing temperatures). The sample was genetically homogeneous[20] and all subjects had the same hair color (black). In the 17th century, only male clan chiefs were buried, whereas after 1700 AD, as many men as women were buried. They were mainly from the elite, and the assumed identity of several subjects and/or their status could be inferred[21] from the associated artifacts, modes of burial and sometimes local legends that specified who was buried in the graves. From the 19th century onwards, with Christianization, almost the entire population was buried and the sample, with the exception of children or some isolated graves, was more equivalent to a random draw in the population[21–23]. At that time people adopted a Russian orthodox way of life and economic wellbeing increased[21].

We studied cases of multiples graves with relatives who died at a similar time. Two women of the same family (n°2 and 6); a woman and one child (n°10 and 11), buried together, had little or no trace of metabolites from tea and tobacco. Two women (n°26 and 12), a mother and daughter[24] who died of smallpox at the same time[25], had different levels of tea and tobacco metabolites; the brother of the daughter (n°29) did not have presence of tea metabolites. Brother and sister were heavy smokers (n°12 and 29) and she was breastfeeding at the time of death.

##### Gender and social status.

Some substances could be associated with special social status *(shaman)* as evidenced by a sub significant association between the status and theobromine concentrations. No significant association was found for gender differences. However, a trend could be confirmed by a greater sample given the much lesser p-values observed after 1800 AD.

| | Estimate | Std. Error | z value | Pr(>\|z\|) |
|---|---|---|---|---|
| **(Intercept)** | -2.121772 | 1.161690 | -1.826 | 0.0678 |
| **Theobromine** | 0.014519 | 0.008582 | 1.692 | 0.0907 |
| **Theophylline** | -0.039655 | 0.047568 | -0.834 | 0.4045 |
| **Caffeine** | -0.007103 | 0.006212 | -1.143 | 0.2528 |
| **Nicotine** | 0.002124 | 0.002538 | 0.837 | 0.4026 |
| **Cotinine** | -0.004708 | 0.005562 | -0.846 | 0.3973 |

*Table S 1. Tea/tobacco use and social status (suspected shamans).*

Tea/tobacco use and gender. No significant association was found for gender differences. However, a trend could be confirmed by a greater sample given the much lesser p-values observed after 1800 AD.

| | | Estimate | Std. Error | z value | Pr(>\|z\|) |
|---|---|---|---|---|---|
| Before 1800 | (Intercept) | -0.0627 | 0.816549 | -0.077 | 0.939 |
| | Theobromine | -0.0004 | 0.004493 | -0.082 | 0.935 |
| | Theophylline | -0.0065 | 0.017675 | -0.370 | 0.712 |
| | Caffeine | 0.0022 | 0.002603 | 0.833 | 0.405 |
| | Nicotine | -0.0052 | 0.006963 | -0.745 | 0.456 |
| | Cotinine | 0.0003 | 0.000664 | 0.397 | 0.691 |
| After 1800 | (Intercept) | -8.1698 | 7.229920 | -1.130 | 0.258 |
| | Theobromine | 0.0769 | 0.062876 | 1.223 | 0.221 |
| | Theophylline | 0.0092 | 0.040560 | 0.227 | 0.821 |
| | Caffeine | -0.0276 | 0.024864 | -1.110 | 0.267 |
| | Nicotine | 0.0310 | 0.029524 | 1.050 | 0.294 |
| | Cotinine | -0.0283 | 0.025701 | -1.102 | 0.271 |

*Table S 2. Tea/tobacco use and gender.*

##### Age classes co-correlations.

Correlation between nicotine and cotinine levels within each of the age class group are given in the table below. Comparison of correlation (co-correlation) between age classes are given (**Erreur ! Source du renvoi introuvable.**) and correlation was found to increase with age. The difference between the young (0-15 years old) and the oldest quartile (over 50 years old) was significant.

| | 0-15 y/o | 15-30 y/o | 30-50 y/o | > 50 y/o |
|---|---|---|---|---|
| **With outliers** | 0.5555 | 0.4529 | 0.4529 | 0.9183 |
| **Without outliers** | 0.5555 | 0.4529 | 0.7316 | -0.1124 |

*Table S 3. Correlation coefficients between Nicotine and Cotinine hair levels amongst age class.*

| | Comparison | N1 / N2 | Fischer's Z | p-value | Zou's C.I. |
|---|---|---|---|---|---|
| With outliers | *0-15 vs 15-30* | 8 / 12 | -0.2711 | 0.7863 | -1.0277 ; 0.6904 |
| | *0-15 vs 30-50* | 8 / 14 | -0.9274 | 0.3537 | -1.2142 ; 0.3326 |
| | *0-15 vs over 50* | 8 / 10 | -2.1887 | 0.0286 | -1.4432 ; -0.0443 |
| Without outliers | *0-15 vs 15-30* | 8 / 12 | -0.2711 | 0.7863 | -1.0277 ; 0.6904 |
| | *0-15 vs 30-50* | 8 / 11 | -1.1145 | 0.2651 | -1.2769 0.2884 |
| | *0-15 vs over 50* | 8 / 9 | 0.6765 | 0.4987 | -0.6761 1.2136 |

*Table S 4. Co-correlation between children and the three other age classes taking or not into account the outliers.*

#### SD 8 Changes in drinking substances and tobacco use over time.

| | | Estimate | Std. Error | z value | Pr(>\|z\|) |
|---|---|---|---|---|---|
| Before 1800 | (Intercept) | -0.0627 | 0.816549 | -0.077 | 0.939 |
| | Theobromine | -0.0004 | 0.004493 | -0.082 | 0.935 |
| | Theophylline | -0.0065 | 0.017675 | -0.370 | 0.712 |
| | Caffeine | 0.0022 | 0.002603 | 0.833 | 0.405 |
| | Nicotine | -0.0052 | 0.006963 | -0.745 | 0.456 |
| | Cotinine | 0.0003 | 0.000664 | 0.397 | 0.691 |
| After 1800 | (Intercept) | -8.1698 | 7.229920 | -1.130 | 0.258 |
| | Theobromine | 0.0769 | 0.062876 | 1.223 | 0.221 |
| | Theophylline | 0.0092 | 0.040560 | 0.227 | 0.821 |
| | Caffeine | -0.0276 | 0.024864 | -1.110 | 0.267 |
| | Nicotine | 0.0310 | 0.029524 | 1.050 | 0.294 |
| | Cotinine | -0.0283 | 0.025701 | -1.102 | 0.271 |

*Table S 5. Changes in drinking substances and tobacco use over time.*

For individuals living before 1800 AD (see Table below), caffeine hair concentration ranged between 11 ng/mg and 252 ng/mg (median 28 ng/mg) while for individuals living after 1800 AD, it ranged between 10 and 9304 (median 55); sub significant association (p-value = 0.0562). No significant association was found for the other xenobiotics (see Table below). Before 1800 AD, some subjects had no tea metabolites, after 1800 AD all had at least one a majority had all three (p = 0.0264). From 1700 to 1750 AD, a majority of individuals had theobromine and caffeine; their number decreased drastically from 1750 to 1800 AD, there were no more after 1800 AD (p = 0.0182). There were no differences between individuals with only theobromine at any time (p = 1).

| | Estimate | Std. Error | z value | Pr(>\|z\|) |
|---|---|---|---|---|
| **(Intercept)** | -0.291535 | 0.608502 | -0.479 | 0.6319 |
| **Theophylline** | -0.038243 | 0.023974 | -1.595 | 0.1107 |
| **Theobromine** | -0.002581 | 0.005238 | -0.493 | 0.6222 |
| **Caffeine** | 0.006761 | 0.003541 | 1.909 | 0.0562 |

*Table S 6. Historical evolution of xenobiotic concentrations.*

| | Estimate | Std. Error | z value | Pr(>\|z\|) |
|---|---|---|---|---|
| **(Intercept)** | -0.7210826 | 0.4288329 | -1.682 | 0.0927 |
| **Nicotine** | 0.0009768 | 0.0015033 | 0.65 | 0.5158 |
| **Cotinine** | -0.0015442 | 0.0019076 | -0.81 | 0.4182 |

#### SD 9 Metabolite concentrations and pipes over geographical range

As for previous analyses, we considered two periods: before and after 1800 AD. For individuals dated before 1800 AD, none of the tests were significant. After 1800 AD, no significant association between geographical regions and metabolite concentration was found using permutation ANOVA (999 permutations, minimum p-value for caffeine: 0.287). However, in the case of caffeine and cotinine, we were able to find a correlation between distance from sampling location to the three main trading posts and/or fairs in the east through which the imported goods had to transit. The highest correlation was with the northeast, which represents the entry into Yakutia of the trade route by which tea was brought at this period from the Pacific harbor of Okhotsk (Table 1 p-value = 0.0112). The robustness of this association was challenged by permuting 10% of the metabolic concentration values among individuals (10000 replicates). Association was confirmed with probability of observing $r_s$ above 0.5 above 0.9 for Zachiversk (Table 2). This led us to suspect a NE-SW gradient in caffeine concentration. No significative association was found between geographic data and any of the other xenobiotics. Substance drinking and tobacco use interpolated over the full range comprised between the archeological sites are shown in Figure S 11 (cf. infra). Before 1800 AD, only isolated individuals showed high levels of derived compounds in hair and after 1800 AD, levels showed a geographic gradient for tea but a gradient was not detectable for tobacco. For pipes, before 1800 AD we were able to find a correlation between distances from people with pipes location to the main closest city, Yakustk (Figure S 12).

| | **Caffeine** | **Theophylline** | **Theobromine** | **Nicotine** | **Cotinine** |
|---|---|---|---|---|---|
| **Yakutsk** | 0.6307 | 0.9071 | 0.5192 | 0.6967 | 0.6705 |
| **Okhotsk** | 0.1252 | 0.7507 | 0.2006 | 0.8799 | 0.4307 |
| **Zachiversk** | 0.0112 | 0.1369 | 0.2589 | 0.5717 | 0.2794 |
| **Verkhoyansk** | 0.0655 | 0.3088 | 0.6500 | 0.1591 | 0.3680 |
| **Kiakhta** | 0.1334 | 0.9336 | 0.3118 | 0.8122 | 0.4384 |
| **Nertchinsk** | 0.1420 | 0.9336 | 0.3403 | 0.8629 | 0.4541 |
| **Verkhnevilyuysk** | 0.1797 | 0.7763 | 0.2006 | 0.6806 | 0.4080 |
| **NE (66.45N, 143.22E)** | 0.0112 | 0.1369 | 0.1509 | 0.5717 | 0.1654 |
| **E (64.17N, 145.13E)** | 0.1097 | 0.7763 | 0.1899 | 0.8629 | 0.4541 |
| **SE (63.57N, 126.50E)** | 0.3403 | 0.9336 | 0.3118 | 0.8290 | 0.5109 |
| **W (62.25N, 116.16E)** | 0.1334 | 0.9336 | 0.3118 | 0.8122 | 0.4384 |
| **NW (66.76N, 123.37E)** | 0.3701 | 0.7763 | 0.1899 | 0.5717 | 0.5621 |

*Table S 7. Absolute correlations between individual xenobiotic hair levels and distance to surrounding cities and four historical gateways to Yakutia.*

For Caffeine and Cotinine, P is the probability of observing $r_s$ above 0.5 among replicates.
For Pipes, P is the probability of observing an association (p-value < 0.05) among replicates

| | *Caffeine* | | *Cotinine* | | *Pipes* | |
|---|---|---|---|---|---|---|
| | $r_s$ | *P* | $r_s$ | *P* | *pval* | *P* |
| **Yakutsk** | 0.1636 | 0.219 | -0.0137 | 0.23 | 0.07 | 0.17 |
| **Okhotsk** | -0.4909 | 0.304 | 0.0410 | 0.219 | 0.60 | 0.00 |
| **Zachiversk** | -0.7273 | 0.931 | -0.3508 | 0.236 | 0.92 | 0.00 |
| **Verkhoyansk** | -0.5727 | 0.886 | -0.3964 | 0.244 | 0.98 | 0.00 |
| **Kiakhta** | 0.4818 | 0.302 | -0.0046 | 0.238 | 0.77 | 0.00 |
| **Nertchinsk** | 0.4727 | 0.298 | -0.0228 | 0.224 | 0.58 | 0.00 |
| **Verkhnevilyuysk** | 0.4364 | 0.264 | 0.0137 | 0.225 | 0.88 | 0.00 |
| **NE (66.45N, 143.22E)** | -0.7273 | 0.923 | -0.3508 | 0.225 | 0.76 | 0.00 |
| **E (64.17N, 145.13E)** | -0.5091 | 0.831 | 0.0592 | 0.208 | 0.30 | 0.00 |
| **SE (63.57N, 126.50E)** | 0.3182 | 0.223 | 0.1230 | 0.216 | 0.08 | 0.15 |
| **W (62.25N, 116.16E)** | 0.4818 | 0.319 | -0.0046 | 0.203 | 0.46 | 0.00 |
| **NW (66.76N, 123.37E)** | 0.3000 | 0.227 | -0.0957 | 0.208 | 0.81 | 0.00 |

*Table S 8. Geography and Caffeine/Cotinine/Pipes: permutation tests (10.000 replicates /10% of permuted data by replicate).*

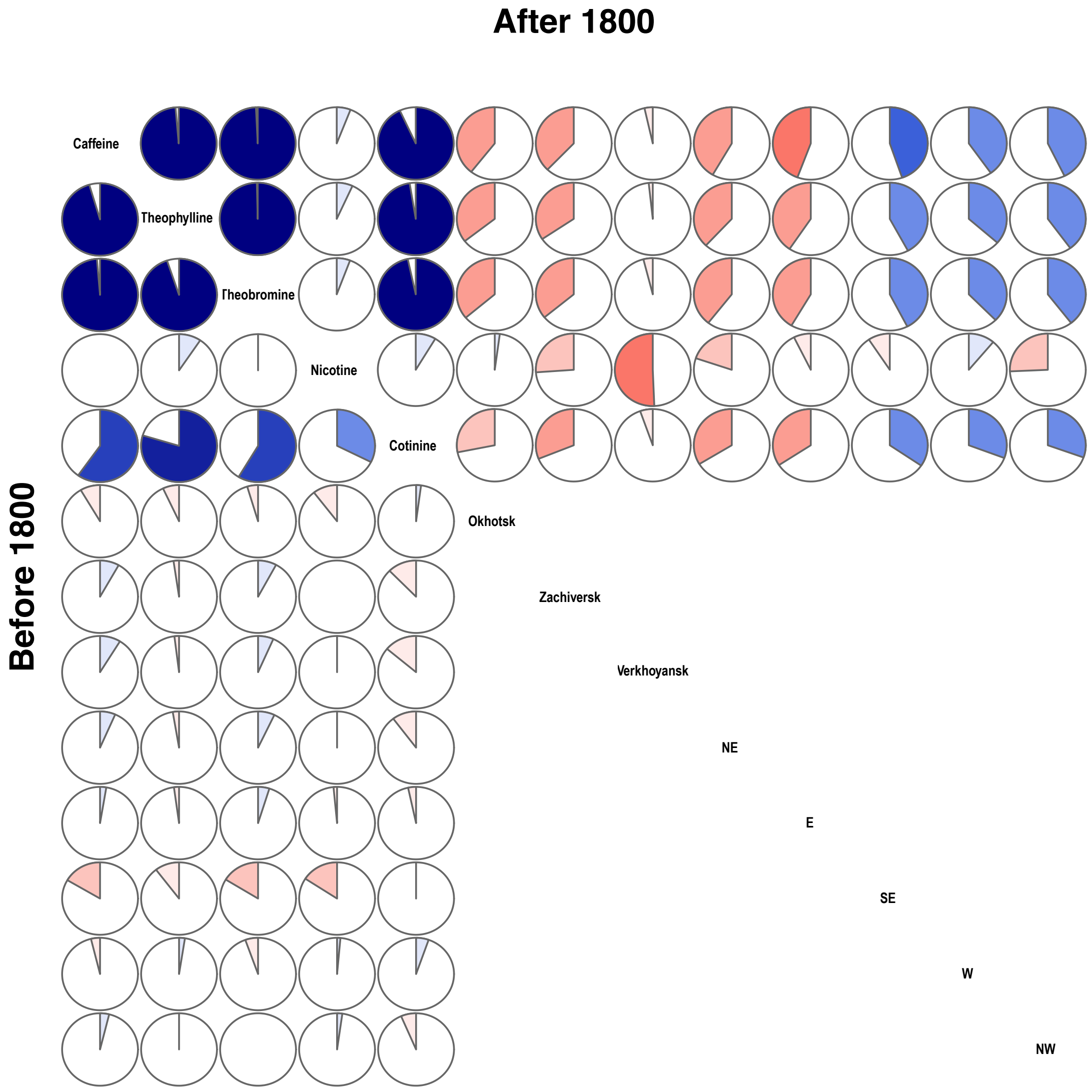

*Figure S 11. Correlogram displaying biochemical/geographical correlations.*

*Upper-diagonal part: before 1800*

*Lower-diagonal part: after 1800*

Positive correlations are shown in blue, negative in red. Color intensity shows absolute strength in correlation. Negative correlations in the east and North means that the further away from these points (distance increases), the more the concentration decreases, resulting in a gradient: dark in the NE > light in the SW.

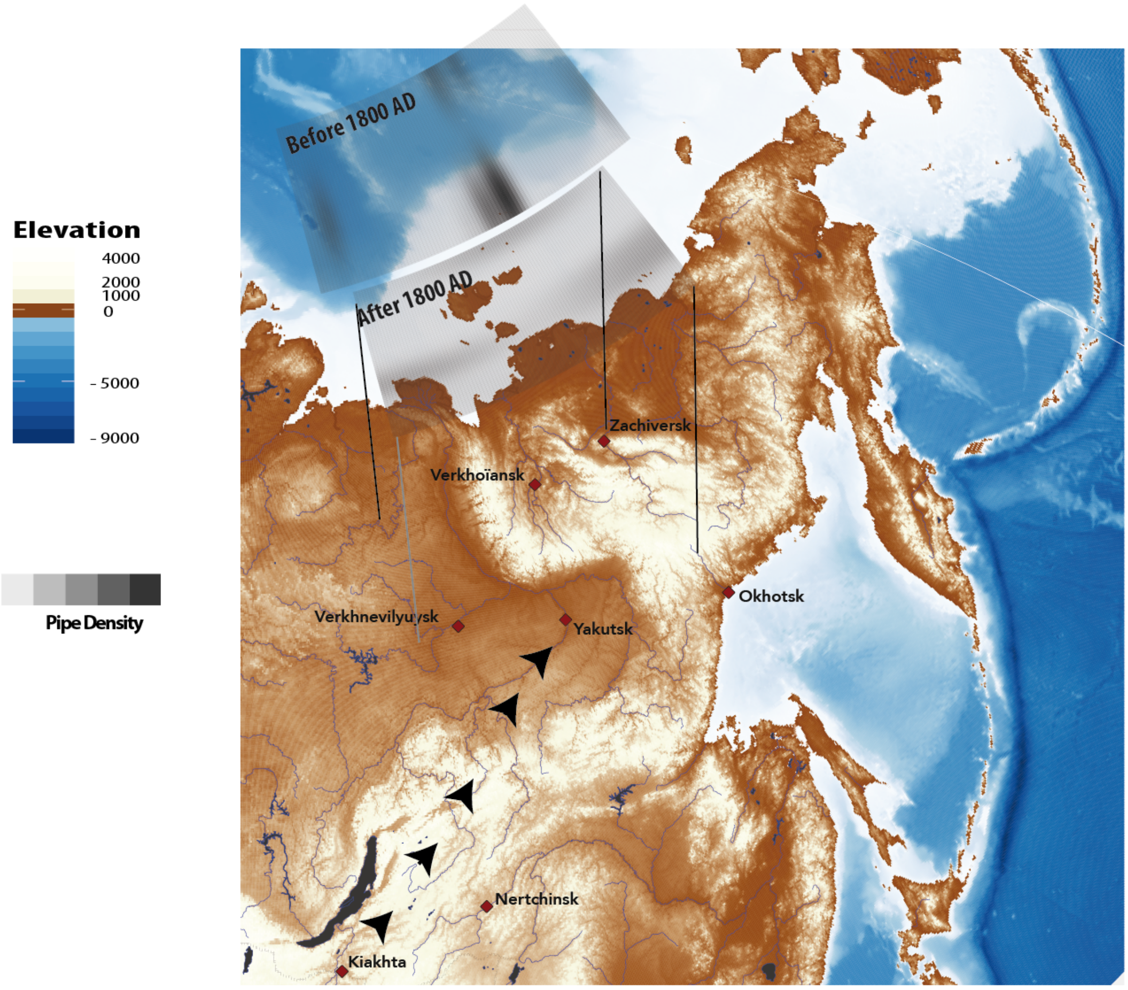

*Figure S 12. Map of eastern Siberia displaying geographical interpolation of pipes found in the graves associated to the bodies with geographical data of the studied area. Before 1800 AD there was a sub-association between pipe ownership and distances to Yakutsk and the South-West of the studied area. The south-west was the route of entry for tobacco from the Kiakhta trading post. After 1800 AD, no geographical pattern was found.*

#### SD 10 Active and/or passive expose

The measurement of nicotine hair concentration can be an informative tool to assess tobacco smoke exposure (whether active or passive) and related illnesses. The main pitfall of nicotine hair concentration interpretation is the impossibility to discriminate between active and passive exposure[26–28]. Indeed, a positive result for nicotine in hair (usually greater than 0.2 ng/mg) could not confirm with any certainty an active use of tobacco by the deceased during their lifetime. Nicotine hair contamination from external sources (such as smoking by other members of the communities in which the deceased individuals lived) cannot be ruled out. Several authors reported the inability of this method to differentiate between active and passive exposure[26–28]. In fact, observed hair concentrations for smokers range usually between 0.4 and 11 ng/mg and between 0.9 and 11 ng/mg for nicotine and cotinine, respectively. But comparable hair values are reported in non-smoker populations, especially in the case of passive exposure[29]. All in all, the determination of nicotine (and/or cotinine) in mummy hair samples cannot definitely confirm active nicotine use during the lifetime of the deceased, as external contamination and exposure to second-hand smoke during their lifetime cannot be excluded[30].

The only case where a pipe was associated with a subject who presented weak traces of cotinine, was with a European imported earthenware pipe - the only one known in Yakutia - which indicates that the pipe was more of a prestige object than an object for smoking (see SD 4).

One of the two subjects with the lowest level of cotinine was a child aged 6 to 12 months and for four subjects where cotinine was detected alone, these levels were low. Present day studies provide subjects with low levels of cotinine not associated with nicotine[31].

Currently, in a majority of individuals, tea and caffeine are present at very high concentrations compared to the two others components[32]. Theophylline and theobromine hair concentrations generally range between 0.1 and 0.6 ng/mg and between 0.3 and 10 ng/mg, respectively[33]. In case of positive result for caffeine, theophylline or theobromine in hair (usually greater than 0.1 ng/mg), there is no data in the literature allowing us to assess the level of tea drinking by the subject.

#### SD 11 Association between level of xenobiotics and biological and /or cultural parameters

Two subjects had only traces of theophylline, three old women had only traces of caffeine and two had traces of both caffeine and theophylline. This could be related to herbal teas that we couldn't identify or to passive exposure to tea. In summer, Yakuts boil water before drinking it and the container used to boil the water may have been used as a teapot beforehand. The possibility - indistinguishable from hair analysis - that three subjects from the 18th century used green and/or black tea and Ivan tea at different times could not be ruled out according to the high level of theobromine compared to theophylline and caffeine levels in their hair. The remains of tea leaf from the teapot tested positive for caffeine (+++), theophylline (++) and theobromine (+), whereas the hair of the woman buried with the teapot tested positive for these three substances but with a concentration of theobromine higher than that of theophylline.

The three women with only low levels of caffeine were three elderly women, two of whom were buried in an area known as Vilyuy. A third was buried in the Indigirka region. It is not impossible that they used a herbal tea based on (or containing) the roots of *Rhodolia Rosea*, which contains caffeine in small quantities[33]. This herbal tea known in the pharmacopoeia of the Siberian people for its anti-fatigue and anti-ageing effect, is still recommended to be taken in large quantities by today's herbalists. This plant grows in the mountains of southern Siberia, but is also known in isolated areas elsewhere, including Villuy[34] where two of the three women were buried.

#### References from SD

1 Crubézy E, Gérard P, Kirianov N, *et al.* The Relationship Between Archaeology, Genetics, Ethnology and History. In: J.-M. Blaising, J. Driessen, J.-P. Legendre and LO, ed. *Clashes of Time*. Louvain: : Presses universitaires de Louvain 2017. 121–38.

2 Zvénigorosky V, Crubézy E, Gibert M, *et al.* The genetics of kinship in remote human groups. *Forensic Sci Int Genet* 2016;**25**:52–62. doi:10.1016/j.fsigen.2016.07.018

3 Mekota A-M, Vermehren M. Determination of optimal rehydration, fixation and staining methods for histological and immunohistochemical analysis of mummified soft tissues. *Biotech Histochem* 2005;**80**:7–13. doi:10.1080/10520290500051146

4 Pichini S, Altieri I, Pellegrini M, *et al.* Hair analysis for nicotine and cotinine: evaluation of extraction procedures, hair treatments, and development of reference material. *Forensic Sci Int* 1997;**84**:243–52. doi:10.1016/S0379-0738(96)02068-3

5 Mahoney GN, Al-Delaimy W. Measurement of nicotine in hair by reversed-phase high-performance liquid chromatography with electrochemical detection. *J Chromatogr B Biomed Sci Appl* 2001;**753**:179–87. doi:10.1016/S0378-4347(00)00540-5

6 Musshoff F, Fels H, Carli A, *et al.* The anatomical mummies of Mombello: detection of cocaine, nicotine, and caffeine in the hair of psychiatric patients of the early 20th century. *Forensic Sci Int* 2017;**270**:20–4. doi:10.1016/j.forsciint.2016.11.011

7 Dulaurent S, Gaulier JM, Imbert L, *et al.* Simultaneous determination of Δ9-tetrahydrocannabinol, cannabidiol, cannabinol and 11-nor-Δ9-tetrahydrocannabinol-9-carboxylic acid in hair using liquid chromatography–tandem mass spectrometry. *Forensic Sci Int* 2014;**236**:151–6. doi:10.1016/j.forsciint.2014.01.004

8 Maublanc J, Dulaurent S, Morichon J, *et al.* Identification and quantification of 35

psychotropic drugs and metabolites in hair by LC-MS/MS: application in forensic toxicology. *Int J Legal Med* 2015;**129**:259–68. doi:10.1007/s00414-014-1005-1
9 Boumrah Y, Humbert L, Phanithavong M, *et al.* In vitro characterization of potential CYP- and UGT-derived metabolites of the psychoactive drug 25B-NBOMe using LC-high resolution MS. *Drug Test Anal* 2016;**8**:248–56. doi:10.1002/dta.1865
10 Kintz P, Richeval C, Jamey C, *et al.* Detection of the designer benzodiazepine metizolam in urine and preliminary data on its metabolism. *Drug Test Anal* 2017;**9**:1026–33. doi:10.1002/dta.2099
11 Wiart J-F, Hakim F, Andry A, *et al.* Pitfalls of toxicological investigations in hair, bones, and nails in extensively decomposed bodies: illustration with two cases. *Int J Legal Med* Published Online First: 6 March 2020. doi:10.1007/s00414-020-02267-3
12 Peters FT, Drummer OH, Musshoff F. Validation of new methods. *Forensic Sci Int* 2007;**165**:216–24. doi:10.1016/j.forsciint.2006.05.021
13 Musshoff F, Madea B. New trends in hair analysis and scientific demands on validation and technical notes. *Forensic Sci Int* 2007;**165**:204–15. doi:10.1016/j.forsciint.2006.05.024
14 Antignac J-P, de Wasch K, Monteau F, *et al.* The ion suppression phenomenon in liquid chromatography–mass spectrometry and its consequences in the field of residue analysis. *Anal Chim Acta* 2005;**529**:129–36. doi:10.1016/j.aca.2004.08.055
15 Zou GY. Toward Using Confidence Intervals to Compare Correlations. *Psychol Methods* 2007;**12**:399–413. doi:10.1037/1082-989X.12.4.399
16 Diedenhofen B, Musch J. Cocor: A comprehensive solution for the statistical comparison of correlations. *PLoS One* 2015;**10**:e0121945. doi:10.1371/journal.pone.0121945
17 Herve M. Testing and Plotting Procedures for Biostatistics “RVAideMemoire”'. CRAN. 2019.https://cran.r-project.org/web/packages/RVAideMemoire/index.html
18 Akima H. Algorithm 761: Scattered-data surface fitting that has the accuracy of a cubic polynomial. *ACM Trans Math Softw* 1996;**22**:362–71. doi:10.1145/232826.232856
19 Singh S, M S, Saini A, *et al.* Breath Carbon Monoxide Levels in Different Forms of Smoking. *Indian J Chest Dis Allied Sci* 2011;**53**:25–8.
20 Crubézy E, Amory S, Keyser C, *et al.* Human evolution in Siberia: from frozen bodies to ancient DNA. *BMC Evol Biol* 2010;**10**:25. doi:10.1186/1471-2148-10-25
21 Crubézy E, Nikolaeva D. *Vainqueurs ou vaincus ?* Paris: : Odile Jacob 2017.
22 Crubézy E, Alexeev A. *Chamane, Kyys jeune fille des glaces*. Paris: : Errance 2007.
23 Crubézy E, Alexeev A. *Мир древних якутов, опыт междисциплинарных сследований. (the world of the ancient Yakuts : a transdisciplinary approach from the French/iakut expedition)*. North East. Yakuts: 2012.
24 Keyser C, Hollard C, Gonzalez A, *et al.* The ancient Yakuts: a population genetic enigma. *Philos Trans R Soc B Biol Sci* 2015;**370**:20130385. doi:10.1098/rstb.2013.0385
25 Thèves C, Biagini P, Crubézy E. The rediscovery of smallpox. *Clin Microbiol Infect* 2014;**20**:210–8. doi:10.1111/1469-0691.12536
26 Haley NJ, Hoffmann D. Analysis of nicotine and cotinine in hair to determine cigarette smoker status. *Clin Chem* 1985;**31**:1598–600.
27 Kintz P. Gas chromatographic analysis of nicotine and cotinine in hair. *J Chromatogr B Biomed Sci Appl* 1992;**580**:347–53. doi:10.1016/0378-4347(92)80542-X
28 Al-Delaimy WK, Crane J, Woodward A. Questionnaire and hair measurement of exposure to tobacco smoke. *J Expo Anal Environ Epidemiol* 2000;**10**:378–84. doi:10.1038/sj.jea.7500102
29 Al-Delaimy WK. Hair as a biomarker for exposure to tobacco smoke. *Tob Control*

2002;**11**:176–82.
30 Musshoff F, Brockmann C, Madea B, *et al.* Ethyl glucuronide findings in hair samples from the mummies of the Capuchin Catacombs of Palermo. *Forensic Sci Int* 2013;**232**:213–7. doi:10.1016/j.forsciint.2013.07.026
31 Li Z, Li Z, Zhang J, *et al.* Using nicotine in scalp hair to assess maternal passive exposure to tobacco smoke. *Environ Pollut* 2017;**222**:276–82. doi:10.1016/j.envpol.2016.12.044
32 Fernández PL, López A, Pablos F, *et al.* The Use of Catechins and Purine Alkaloids as Descriptors for the Differentiation of Tea Beverages. *Microchim Acta* 2003;**142**:79–84. doi:10.1007/s00604-003-0002-8
33 Tracqui A, Kintz P, Mangin P. Hair analysis: a worthless tool for therapeutic compliance monitoring. *Forensic Sci Int* 1995;**70**:183–9. doi:10.1016/0379-0738(94)01626-G
34 Pozzhim A V, P SY, A. KG. Rhodiola in Southern Siberia. In: *Areals of plants of the flora of the USSR. Op. 3. L (In Russian)*. Leningrad: : Izd. Leningr. un-ta 1976.
